# Leveraging pleiotropy identifies common-variant associations with selective IgA deficiency

**DOI:** 10.1101/2024.06.24.24309378

**Authors:** Thomas W. Willis, Effrossyni Gkrania-Klotsas, Nicholas J. Wareham, Eoin F. McKinney, Paul A. Lyons, Kenneth G.C. Smith, Chris Wallace

## Abstract

Selective IgA deficiency (SIgAD) is the most common inborn error of immunity (IEI). Unlike many IEIs, evidence of a role for highly penetrant rare variants in SIgAD is lacking. Known SIgAD-associated variants are common in the general population, but previous studies have had limited power to identify common-variant associations due to their small sample size. We sought to overcome this problem first through meta-analysis of two existing GWAS. This identified four novel common-variant associations and we found also that SIgAD-associated variants were enriched in genes known to harbour variants causal for Mendelian IEIs. SIgAD showed evidence of shared genetic architecture with serum IgA and a number of immune-mediated diseases. To further enhance power, we leveraged this pleiotropy through the conditional false discovery rate procedure, conditioning our SIgAD meta-analysis on large GWAS of asthma and rheumatoid arthritis, and our own meta-analysis of serum IgA. This identified an additional 17 variants associated with SIgAD. Our results increase the number of known SIgAD-associated variants outside the MHC to 26 and strengthen the evidence for a polygenic, common-variant aetiology for SIgAD, highlighting both T- and B-cell biology in the development of this disease. Our approach to genetic variant discovery is relevant to the study of other rare diseases and we hypothesise genes newly associated with SIgAD might be explored for as-yet elusive rare-variant associations with SIgAD or IEIs more generally.

## 1. Introduction

Inborn errors of immunity (IEIs) comprise a group of 485 diseases characterised by immune dysfunction of genetic origin [1]. Selective IgA deficiency (SIgAD) is the most common of these disorders in Europeans. SIgAD is defined by a serum IgA level below 7mg/dL in individuals four years and older in whom serum IgG and IgM are normal, and for whom other causes of hypogammaglobulinaemia have been excluded [2]; some authors also allow for the presence of IgG subclass deficiency in SIgAD [3,4]. SIgAD has a prevalence in individuals of European ancestry between 1:300 and 1:1,200 [5]. Higher estimates have been reported in Saudi Arabia (1:143) [6] and Nigeria (1:252) [7], but the disorder appears to be less common in East Asian ancestries such as Japanese (1:14,840) [8] and Han Chinese (1:3,230) [9].

The fundamental defect in SIgAD is a deficiency in IgA production. IgA is the most abundant antibody in the body [10] and exists in monomeric and mainly dimeric forms in the serum and on the mucosae, respectively; other quaternary structures of secretory IgA, including trimers and tetramers, are known [11]. IgA is recognised as the principal antibody isotype in mucosal immunity in the respiratory, gastrointestinal, and genitourinary tracts [12]. The distinct role of serum IgA remains somewhat obscure [13]. Secretory IgA effects immune exclusion of pathogens at mucosal surfaces by binding to cell-surface microbial antigens required for pathogen adherence to epithelia and thereby blocking their interaction with host ligands, as well as sterically hindering pathogen transit [14]. IgA’s binding to cell-surface antigens can also entrap microbes in the mucus for removal by peristalsis or the mucociliary escalator, whilst binding of soluble virulence factors such as toxins can act to ‘quench’ microbial virulence [14]. IgA also mediates homeostasis of the intestinal microbiome [15].

There is not yet a conclusive account of SIgAD’s pathogenesis but the underlying immune defects include failures of B-cell maturation, class switch recombination, and differentiation into plasma cells and memory B cells [16,17]. If IgA deficiency manifests clinically, it does so most often as recurrent sinopulmonary infections, the prevalence of which was recently estimated as 51% in SIgAD patients [18]. Gastrointestinal infection is less common (16%) [18]; that immunodeficiency more often manifests in the respiratory tract than in the gastrointestinal tract may relate to the compensatory production of secretory IgM in the latter [19]. Broader immune dysfunction in the form of allergic and autoimmune disease are present in 29% and 22% of patients, respectively [18].

As many as three-quarters of SIgAD cases may be asymptomatic [20]. A SIgAD prevalence of 1 in 328 was reported in a study of healthy American blood donors [21]. A compensatory increase in IgM and IgG secretion may explain the ability of IgA-deficient individuals to remain healthy [20].

The variation in SIgAD’s prevalence by ancestry and its pattern of familial aggregation [22–24] are adduced as evidence of a genetic origin. Deletions of the short [25,26] and long [27–29] arms of chromosome 18 and the presence of a ring-18 chromosome [30] were the first genetic defects linked with IgA deficiency but these have not led to the identification of candidate genes [31,32]. Later studies identified the major histocompatibility complex (MHC) as a SIgAD susceptibility locus, with some dispute over whether this signal should be mapped to the class II or III regions [33–36]. The 8.1 ancestral MHC haplotype (8.1 AH) was identified as the ‘single strongest genetic risk factor’ for SIgAD in Northern Europeans [37–39]. The only SNP fine-mapping study of the MHC in SIgAD identified four independent risk and protective effects in the class II and III regions [39].

In contrast with many IEIs, evidence of highly penetrant monogenic mutations in SIgAD is lacking [40]. An association of common variable immunodeficiency (CVID) and SIgAD with mutations of incomplete penetrance in *TNFRSF13B* has been reported [41–44], but this finding could not be replicated in a larger SIgAD cohort [45]. IgA deficiency can occur as part of rare syndromic genetic disorders with better-defined genetic aetiologies [40] which are definitionally excluded from SIgAD. For example, IgA deficiency may feature in the autosomal recessive disorder ataxia telangiectasia [46], which is caused by loss-of-function mutations in *ATM*.

Two genome-wide association studies (GWAS) conducted in overlapping European cohorts identified SIgAD associations with common variants mapping to the genes *IFIH1*, *CLEC16A*, *PVT1*, and *AHI1*, and the *ATG13-AMBRA1* locus [47,48]. The multiplicity of common variant associations identified thus far and the absence of a clear monogenic aetiology are suggestive of a polygenic origin for SIgAD.

Power to discover common SIgAD-associated variants, particularly with small to moderate effect sizes, has been limited by the small size of study cohorts, a problem inherent to the study of rare diseases more generally. SIgAD’s putative genetic origin suggests there remains considerable scope for the discovery of more common variants. Here we aimed to overcome the small sample limitation through the use of meta-analysis and the pleiotropy-informed conditional false discovery rate (cFDR) [49,50]. The first increases sample size while the second leverages information from GWAS of related traits, with both techniques designed to increase power. We first perform a meta-analysis of two existing SIgAD GWAS, identifying four novel associations. We perform an additional meta-analysis of serum IgA production in the general population as a trait ostensibly related to SIgAD, increasing the sample size over the largest existing study of this SIgAD-related trait from 41,448 to 57,063. We thereby identify eight novel serum IgA associations. We then leverage these IgA data and two additional traits genetically related to SIgAD to identify 17 novel SIgAD-associated variants. Many of these variants map to immunologically salient genes, including some implicated in Mendelian IEIs.

## 2. Material and methods

### 2.1 GWAS data sets and their preprocessing

We made use of GWAS data sets in four aspects of our work. We first meta-analysed two publicly available SIgAD studies by Bronson and colleagues [48] and the FinnGen consortium [51]. We supplemented this with an analysis of serum IgA measured in healthy individuals, meta-analysing our own novel study of 7,938 subjects with two other GWAS by Liu et al. [52] and Dennis et al. [53]. We then estimated SIgAD’s genetic correlation with a panel of immune-mediated diseases: asthma [54], Crohn’s disease and ulcerative colitis [55], juvenile idiopathic arthritis [56], multiple sclerosis [57], primary biliary cholangitis [58], primary sclerosing cholangitis [59], rheumatoid arthritis [60], systemic lupus erythematosus [61], type 1 diabetes [62], dermatitis and eczema [51], hyperparathyroidism [51], hypothyroidism [51], Addison’s disease [63], and IgA nephropathy [64]. Lastly, we leveraged our meta-analysis of serum IgA, and published GWAS of asthma [54] and rheumatoid arthritis [65] in a cFDR analysis of our meta-analysis of SIgAD.

We tabulate these GWAS data sets in Supplementary Table 1. The majority of GWAS data sets used were publicly available and were downloaded from the EBI GWAS Catalog [66], FinnGen [51] and the Pan-UK Biobank data repositories, [54], or bespoke websites in the case of the IgA nephropathy [64] and rheumatoid arthritis [65] data sets. Where possible, we give URLs to download these data sets (Supplementary Table 1). The multiple sclerosis GWAS data set is not publicly available but can be requested as the ‘MS Chip (Science 2019)’ data set from the IMSGC webpage linked in Supplementary Table 1.

The GWAS data sets used in all analyses were preprocessed into a uniform format with the *GWAS_tools* pipeline (https://github.com/GRealesM/GWAS_tools) to facilitate joint analysis; we used a fork of the pipeline customised for our purposes (https://github.com/twillis209/GWAS_tools). Missing effect estimates and standard errors were recomputed from odds ratios and p-values where necessary. We lifted over genomic coordinates in data sets using the earlier hg19 genome assembly to hg38 with the UCSC *liftOver* executable [67]. We then merged each data set with a panel of SNPs taken from the 1000 Genomes Phase 3 (1kGP3) high-coverage data set [68]; we processed the 1kGP3 data set to contain only SNPs with MAF > 0.005 in European individuals, as we sought to investigate common and low-frequency variant architecture. After establishing that alleles in all data sets were stated relative to the positive strand, we aligned all data sets to the allele order in the 1kGP3 panel. We encountered an apparent mislabelling of the ‘effect’ and ‘other’ allele columns in the Bronson data set available on the EBI GWAS Catalog. We resolved this by cross-referencing the alleles and effects reported in their paper and the GWAS Catalog’s lead SNP summary (Supplementary methods).

Bronson and colleagues confined their SIgAD cohort to individuals ‘without comorbid autoimmune diseases’ including coeliac disease [48]. The Bronson cohort overlaps with that recruited by Ferreira and colleagues for their earlier SIgAD GWAS [47], the cohort for which contributed 760 cases and 1,724 controls to the Bronson cohort. The Ferreira SIgAD case cohort included individuals referred for study inclusion due to ‘infection proneness’ and in whom IgA deficiency was confirmed by serum analysis. It also included blood donors whose IgA deficiency was identified incidentally during screening of blood donations; some of these IgA-deficient blood donors did also report infection proneness.

The FinnGen cohort did not exclude patients on the basis of autoimmunity and there exists case overlap with autoimmune diseases. SIgAD case overlap with other phenotypes is tabulated under the ‘Correlations’ heading on the Risteys platform in terms of the Jaccard index: https://r10.risteys.finngen.fi/endpoints/D3_DEF_IIGA.

### 2.2 GWAS of serum IgA

Participants were recruited from the European Prospective Investigation into Cancer (EPIC)-Norfolk Study cohort (https://doi.org/10.22025/2019.10.105.00004). The study was carried out according to the principles of the Declaration of Helsinki, participants provided informed consent, and the study was approved by the Norwich Local Research Ethics Committee.

Measurement of IgA alongside IgG1, IgG2, IgG3, IgG4, and IgM was performed for 9,941 participants using the Prototype Human Isotyping 6-plex ElectroChemiLuminescent Immunosorbent Assay (Meso Scale Discovery, USA) and read using the MSD Sector Imager 6000 instrument. All incubation was at room temperature, sealed, 600rpm, 3mm orbital shaking. All plate washing was performed with 3×300µL/well 1x PBS-T (New England Biolabs, UK) on the BioTek 405LS Microplate Washer and then blotted dry. Plates were prepared on the Tecan Fluent 780. Plates were incubated for one hour with 150µL/well of MSD Blocker A solution (Meso Scale Discovery, USA). Eight standards were prepared by spiking Diluent 100 with the MSD Calibrator Stocks for each of the analytes (including one blank standard). After washing, 25µL/well of sample was added and plates incubated for two hours; pooled serum samples and unknown samples were diluted 1:100000 in Diluent 100 (Meso Scale Discovery, USA) and standards were undiluted. After washing, 25µL/well of 1x MSD Detection Antibody Solution was added and plates incubated for 2 hours. Plates were finally washed and read with 150µL/well 2x MSD Read Buffer T with Surfactant (Meso Scale Discovery, USA). Interpolation of the Standard Curve was performed using MSD Discovery Workbench software using a 4-PL fitting algorithm with 1/y^2^ weighting on a log-log scale.

The following quality control parameters were applied:

1. calculated concentration of standards and pooled serum samples should be within ±20% from the mean for at least 50% of individual replicates in each sample group
2. The coefficient of variation for standards, pooled serum samples, and unknown samples should be ≤20% for at least 50% of individual replicates in each sample group (excluding samples outside of the detection range)
3. signal coefficient of variation of standards should be ≤20% for at least 50% of individual replicates
4. The distribution parameters of plates should be ±30% from the overall value for at least two of the 25th, 50th and 75th percentiles; the 2.5th and 97.5th percentiles were monitored.

9,611 samples returned valid measurements for IgA after applying these quality control parameters.

Participants were genotyped using the Affymetrix UK Biobank Axiom array. Samples were excluded for the following reasons: failed channel contrast (DishQC <0.82); low call rate (<0.97); gender mismatch between reported sex and genetic sex; outlying heterozygosity; unusually high number of singleton genotypes; or impossible identity-by-descent values. SNPs were removed if they met one of the following conditions: call rate < 0.95; clusters failed Affymetrix SNPolisher standard tests and thresholds; MAF was significantly affected by plate; SNP was a duplicate based on chromosome, position and alleles (selecting the best probeset according to Affymetrix SNPolisher); p < 10^-6^ for a test of Hardy-Weinberg equilibrium; alleles did not match the reference; or MAF = 0.

Prior to running the GWAS, samples were removed if they had: age greater than 80 years at the time of sampling; absence of genetic data; non-European ancestry; presence of relatives in the EPIC Norfolk sample as indicated by *π* >= 0.1875. Variants with imputation quality (info) < 0.4, Hardy-Weinberg Equilibrium p-value < 1×10^-6^, minor allele frequency (MAF) < 0.001, or effect size/standard error > 10 were removed. The log-transformed IgA phenotypes were standardised and the GWAS was performed using an additive model in SNPTEST (v2.5.4-beta3) incorporating age, sex, and scores on the first ten principal components of the genetic relatedness matrix.

### 2.3 GWAS meta-analysis

For both the SIgAD and serum IgA phenotypes we conducted a fixed-effect, inverse variance-weighted meta-analysis of GWAS summary statistics. We performed a left join of the SNPs in the Bronson data set onto the FinnGen data set, retaining only those FinnGen SNPs which were also present in the Bronson data set as the latter was the more highly-powered study. For the serum IgA data sets, we performed a left join of the SNPs in the Liu data set onto those in our own data set and the Dennis data set, retaining only those present in Liu as the more highly-powered study.

### 2.4 Enrichment testing

We tested whether our SIgAD GWAS signals were enriched in genes known to harbour rare genetic causes of IEIs. We obtained the latest IUIS-curated list of IEIs and their causal genetic defects [1]. Exclusion of chromosomal aberrations left 448 genes known to harbour variants causal for monogenic IEIs (Supplementary Data 1). We retrieved the coordinates of these genes using the Open Targets Genetics [69,70] and Ensembl [71] APIs, and added 50kb flank regions up- and downstream of each gene. We identified the subset of SNPs from our GWAS meta-analysis which fell inside these genic and perigenic regions and computed the empirical 50th, 90th, 95th, and 99th percentiles of their test statistics. To test the null hypothesis that there was no difference in the enrichment of association signals between SNPs in (peri-)genic IEI regions and SNPs in a comparable pan-gene set, we repeatedly sampled 448 genes from the set of all genes and computed the aforementioned percentiles in each sample. From 5,000 realisations, we obtained a p-value as the proportion of samples with an order statistic (50th, 90th, 95th, and 99th) equal to or exceeding that observed in the data.

### 2.5 Genetic correlation estimation

We estimated the genetic correlation between SIgAD, serum IgA, and a selection of IMDs using LDAK version 5.2 (https://dougspeed.com/ldak). We generated SNP taggings using the 1kGP3 data described above. We excluded SNPs which explained more than 1% of the phenotypic variance, as per the LDAK authors’ recommendation, as analyses can be sensitive to large-effect loci.

To estimate the genetic correlation, we performed an inner join of each pair of data sets and removed an extended interval from chromosome six containing the MHC (chromosome 6, positions 24,000,000 to 45,000,000) as the locus’s strong, long-range patterns of linkage disequilibrium (LD) are not representative of those observed genome-wide. We used the ‘LDAK-Thin’ heritability model when generating taggings and the default ‘SumHer-GC’ model of confounding, which assumes that confounding inflation is multiplicative (http://dougspeed.com/genetic-correlations). We performed a Wald test against a null hypothesis of no genetic correlation (i.e. H_0_: r_g_ = 0). We used the *stats::p.adjust* function in R to control the FDR at 0.05 across these chi-squared tests using the Benjamini-Hochberg method [72].

### 2.6 The GPS test

The genetic correlation measures the average effect of pleiotropy across the genome [73] and is comparatively underpowered for small samples [74]. We supplemented it with the genome-wide pairwise-association signal sharing (GPS) test as the statistic of the latter takes the supremum of a weighted norm and has greater power than a test of the genetic correlation in small samples [75]. We used our own implementation of the (GPS) test (https://github.com/twillis209/gps_cpp) which is designed to detect genetic overlap in the small sample context [75,76]. For each pair of GWAS data sets tested we first performed an inner join of the data sets and removed the MHC as described above. We then subjected the remaining SNPs to linkage disequilibrium (LD) pruning, which removed a single SNP from each SNP pair such that the remaining SNPs satisfied the constraint r^2^ < 0.8. We used the European 1kGP3 panel discussed above to compute pairwise r^2^ estimates. r^2^ estimation and LD pruning was carried out using *PLINK v2.00a6LM* [77,78].

### 2.7 Mendelian randomisation

We carried out two-sample Mendelian randomisation to assess the causal effect of serum IgA, an exposure, on the risk of two IgA-related disease outcomes: SIgAD and IgA nephropathy (IgAN). We selected as our genetic instruments the lead SNPs from the genome-wide significant loci identified in our meta-analysis of serum IgA (Supplementary Table 2). We performed an inverse variance-weighted analysis using version 0.90 of the R package *MendelianRandomisation* [79]; we give the input SNP effect sizes and standard errors for IgA, SIgAD, and IgAN in Supplementary Table 3.

An assumption of Mendelian randomisation is that the instrumental variables’ effect on the outcome is mediated solely by their effect on the exposure, i.e. they do not exhibit horizontal pleiotropy [80]. We performed colocalisation analysis of each IgA-associated SNP with SIgAD and IgAN to identify loci at which there may be distinct causal variants: a causal variant distinct from that underlying the IgA association and independently associated with the disease outcome would suggest an effect on the outcome not mediated by the exposure, a violation of the assumption just stated. On the basis of these analyses we excluded a single SNP each from the IgA-SIgAD and IgA-IgAN data sets: rs16830188 and rs3181356, respectively. We performed colocalisation analyses with version 5.2.3 of the R package *coloc* using the *coloc.abf* function with default priors. We used a 100-kilobase window around each SNP to define the set of SNPs used in each analysis.

### 2.8 Conditional false discovery rate analysis

GWAS conducted in small cohorts lack power to detect all but the strongest associations with a trait. Where a trait shows evidence of shared genetic architecture with other traits for which larger GWAS exist, we can use the cFDR procedure to leverage information from these related traits, thereby increasing power to detect associations whilst controlling the type 1 error rate [81]. The cFDR conditions p-values relating to the association of SNPs with the ‘principal’ trait of interest on p-values relating to the association of the same SNPs with related ‘auxiliary’ traits. This conditioning procedure leverages evidence of the association of a SNP with these related auxiliary traits as evidence for association of the same SNP with the principal trait; the principal p-value is modulated in accordance with this evidence to produce a pleiotropy-informed p-value, the ‘v-value’. We used the cFDR procedure to condition p-values of SNP association with SIgAD on p-values from GWAS of asthma and rheumatoid arthritis, and our own meta-analysis of serum IgA, hoping to improve power to discover SIgAD-associated SNPs.

To carry out the cFDR procedure, we used our own fork of the *cfdr* package [50] which implements the computationally intensive *vl* function in C++ rather than in less performant R code (https://github.com/twillis209/cfdr). We performed a left join of each of our three chosen conditioning data sets onto the SIgAD meta-analysis data set and removed the MHC from the merged data set as described above. For each of the three SIgAD-conditioning data set pairs, we performed LD pruning as described above but imposed the constraint r^2^ < 0.2 to provide a more independent set for maximum likelihood estimation of the parameters of the joint null distribution P, Q | H^p^_0_ using the *cfdr::fit.2g function*. A key assumption of the cFDR method is that the p-values of the principal trait (here SIgAD) are uniformly distributed under the null hypothesis; we enforced this by applying a genomic control correction to the principal trait’s test statistics. We performed a three-step iterative cFDR analysis on rheumatoid arthritis, asthma, and serum IgA, carrying forward the v-values (the modulated p-values which constitute the cFDR procedure’s output) by the preceding step for use as the principal p-values in the second and third iterations. Where auxiliary p-values were absent in an iteration due to the absence of SNPs in a conditioning data set, we carried forward an unmodified principal p- or v-value from the previous iteration.

Control of type 1 error may be lost in a cFDR analysis when iteratively conditioning the principal trait on multiple auxiliary covariates displaying ‘extreme’ dependence [81]. This motivated our choice of three iterations.

## 3 Results

### 3.1 A small increase in sample size yields four novel SIgAD associations

Our meta-analysis of two SIgAD GWAS by Bronson and colleagues [48] and FinnGen [51] achieved an 8% increase in the number of cases over the previous largest study (from 1,635 to 1,761; Supplementary Table 1). Notably, we identified four novel associations among the nine genome-wide significant associations (Table 1 and Figure 1).

**Figure 1.**
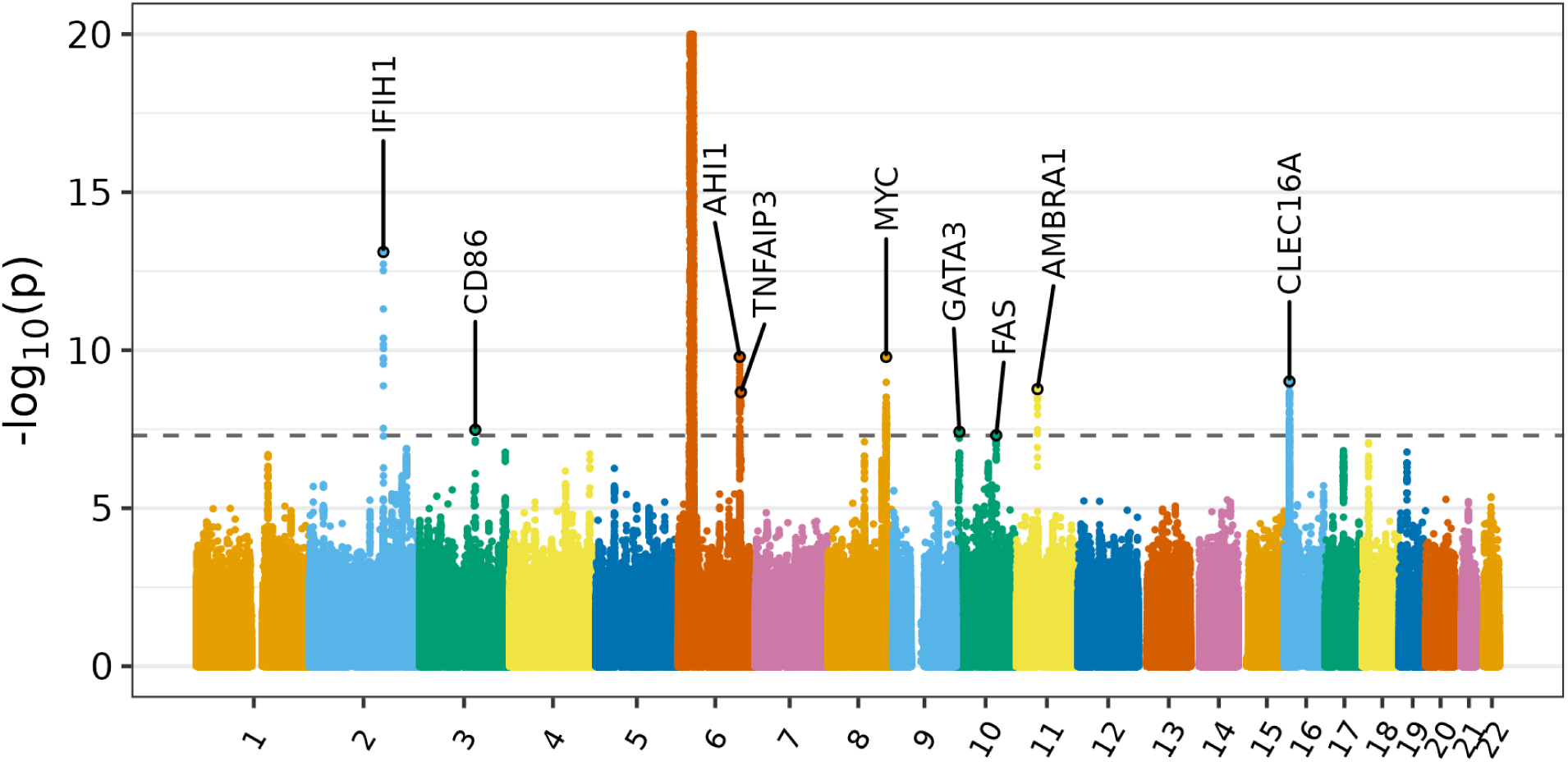
Manhattan plot showing results of GWAS meta analysis of SIgAD. Association signals are labelled with the genes to which they were mapped based on proximity and functional data. The dashed line indicates the threshold of genome-wide significance, p < 5×10^-8^. The signal in the MHC on chromosome 6 is truncated at 20.

**Table 1.** Lead SNPs from genome-wide significant associations in the SIgAD GWAS meta-analysis. The ‘Variant’ column gives the rsID of each SNP, and the reference and effect alleles separated by ‘>’’. EAF is effect allele frequency. Effect allele frequencies given were obtained from gnomAD’s estimate in non-Finnish Europeans. ‘Gene’ gives the gene(s) with the most evidence linking it/them to the association signal. ‘Novel’ indicates whether an association with SIgAD has previously been reported for a SNP. ‘IEI gene’ indicates whether the SNP is located in, near, or is otherwise associated with a gene known to harbour variants causal for IEIs. ‘OR’ is odds ratio.

| Variant | Chromosome | Position | EAF | Gene | Novel | IEI gene | OR | p-value |
| --- | --- | --- | --- | --- | --- | --- | --- | --- |
| rs1990760:C>T | 2 | 162,267,541 | 0.62 | *IFIH1* |  |  | 1.38 | 7.78E-14 |
| rs9831894:A>C | 3 | 122,081,640 | 0.40 | *CD86* | TRUE |  | 1.26 | 3.30E-08 |
| rs2179781:C>A | 6 | 135,398,362 | 0.55 | *AHI1* |  |  | 0.77 | 1.62E-10 |
| rs112920346:C>T | 6 | 137,833,918 | 0.14 | *TNFAIP3* | TRUE | TRUE | 0.71 | 2.12E-09 |
| rs72722767:G>A | 8 | 128,192,857 | 0.29 | *MYC* |  |  | 0.75 | 1.64E-10 |
| rs1244181:A>G | 10 | 8,049,414 | 0.82 | *GATA3* | TRUE |  | 1.35 | 3.83E-08 |
| rs2031613:C>T | 10 | 89,007,167 | 0.71 | *FAS* | TRUE | TRUE | 0.79 | 4.93E-08 |
| rs61882719:A>G | 11 | 46,517,392 | 0.30 | *AMBRA1* |  |  | 1.35 | 1.70E-09 |
| rs34443974:C>T | 16 | 11,085,448 | 0.20 | *CLEC16A* |  |  | 0.72 | 9.71E-10 |

The lead SNP for the first of these novel associations, rs112920346, occurred as one of nine significant common SNPs in a signal centered on *TNFAIP3* (Supplementary Figure 1). This gene encodes tumour necrosis factor α-induced protein 3 (also known as A20), a deubiquitinating enzyme and key negative regulator of NF-κB [82], which is in turn a critical mediator of inflammatory responses in both innate and adaptive immunity [83]. Common variants mapped to *TNFAIP3* are associated with a range of immune-mediated diseases including systemic lupus erythematosus [84], rheumatoid arthritis [65], psoriasis [85], multiple sclerosis [86], and Sjoegren’s syndrome [87]. We confirmed that the *AHI1* and *TNFAIP3* signals were distinct with r^2^ = 0.001 between lead SNPs (Supplementary Figure 2).

*TNFAIP3* is the most obvious candidate for the gene mediating this signal’s effect as a protein-coding gene harbouring IEI-causal variants, but the *WAKMAR2* lncRNA gene overlaps the promoter and gene body of *TNFAIP3* [88], and has been identified as an antisense ‘enhancer RNA’ gene whose transcription into a lncRNA may mediate its effects on gene expression [89]. Its lncRNA gene product downregulates the production of inflammatory chemokines by keratinocytes during wound healing [88], is itself downregulated in the lesional skin of psoriasis patients [88,90], and was shown to regulate a range of immune-related genes in invasive breast cancer, including the interleukin receptor subunit gene *IL27RA* and the immunoglobulin A and D constant region heavy chain genes *IGHA1* and *IGHD* [89]. However, *WAKMAR2* knockdown does not affect the level of TNFAIP3 in keratinocytes, suggesting it is not an antisense RNA for *TNFAIP3* in particular[88].

We identified a second novel association with a SNP in an intron of another TNF- and IEI-related gene, *FAS*, which encodes the cell-surface death receptor Fas. The lead SNP rs2031613 is a *cis*-eQTL for *FAS* and *ACTA2* in several tissues including T cells [91]. We identified a third novel association with rs1244181, a SNP which lies upstream of the gene *GATA3* and is associated with lymphocyte counts [92,93]. *GATA3* is a transcription factor and lymphocyte lineage specifier which promotes T cell specification over B cell potential in T cell precursors [94].

The final novel association signal we identified had its lead SNP in an intron of *CD86* (Supplementary Figure 3). As of the latest IUIS report on IEIs [1], *CD86* is not known to harbour variants causal for any Mendelian IEI.

### 3.2 Regions containing IEI genes are enriched for polygenic signal

We linked two novel SIgAD GWAS signals to genes harbouring highly penetrant variants causal for monogenic IEIs (Supplementary Table 4). Rare loss-of-function mutations in *TNFAIP3* cause the autosomal dominant IEI familial Behçet-like autoinflammatory syndrome-1, an autoinflammatory disorder characterised by systemic inflammation, mucosal ulceration, polyarthritis, uveitis, and recurrent infections in some [95,96]. Similarly, mutations in *FAS* cause autoimmune lymphoproliferative syndrome (ALPS), an IEI and disease of immune dysregulation [1,97]. Fas dysfunction in ALPS leads to defective apoptosis, of which the most salient manifestation is chronic lymphoproliferation, and impaired B cell selection in the germinal centre which in turn leads to loss of peripheral tolerance and the production of autoantibodies [98,99]. To investigate whether these associations reflected a general tendency for IEI genes to harbour common SIgAD-associated variants, we examined whether SNPs lying in or near genes known to harbour rare variants which cause Mendelian IEIs showed more evidence of association with SIgAD than did SNPs in comparable regions centered on genes in general. We found significant enrichment of association signals at and above the 90th percentile (p < 0.0005) in IEI-proximate genic and perigenic SNPs using the test statistics of our SIgAD meta-analysis (Figure 2).

**Figure 2.**
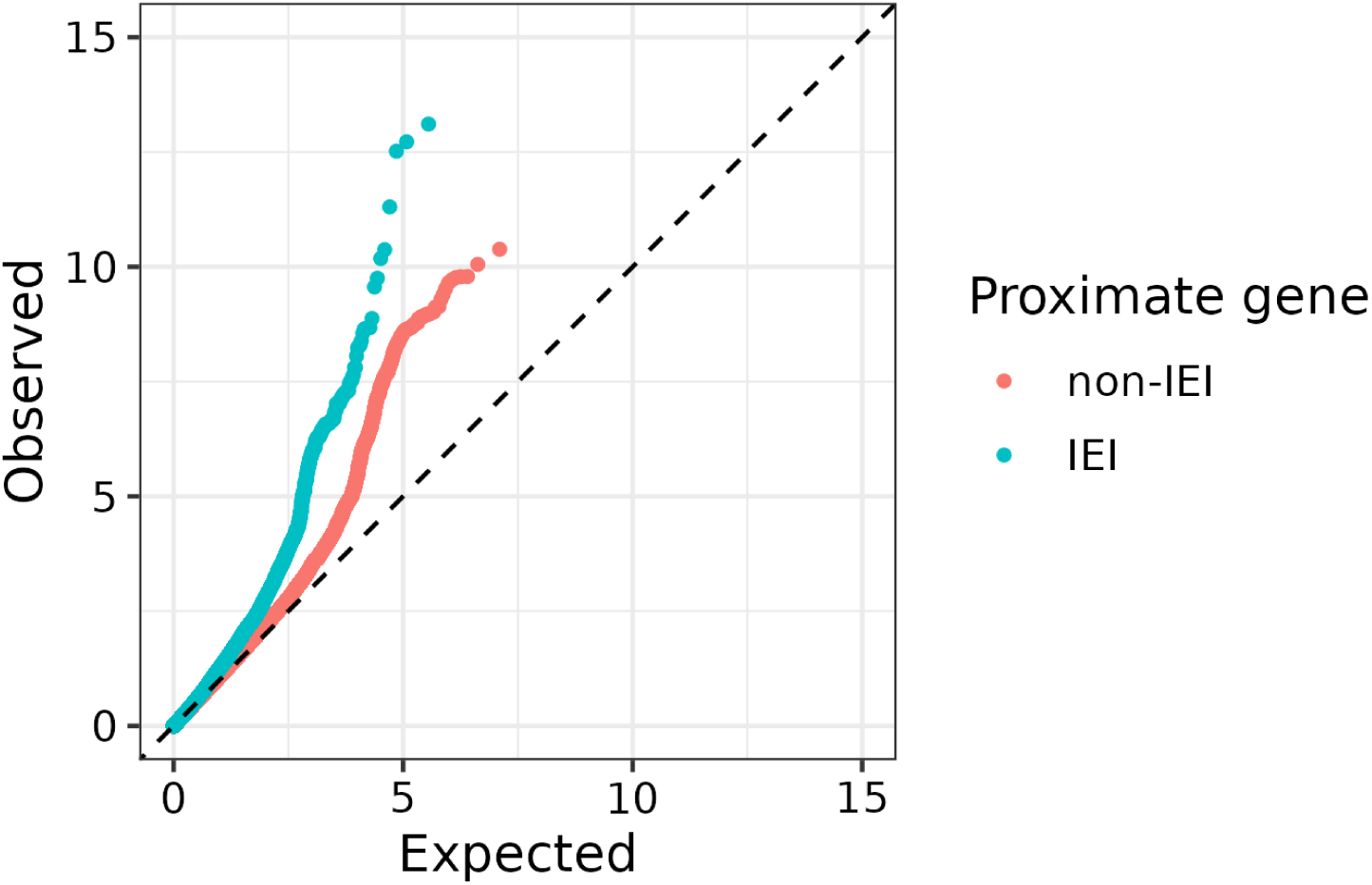
A quantile-quantile plot depicting the empirical quantiles of the SIgAD meta-analysis p-values on a -log10 scale, coloured according to their proximity to IEI or non-IEI genes. Each point corresponds to a SNP, with the y-coordinate giving the SNP’s observed -log10 p-value in the meta-analysis and the x-coordinate giving the -log10 p-value expected if there were no associated SNPs. The dotted line y = x depicts the trajectory of the data where the observed distribution matches the expected uniform distribution, i.e. the scenario where all SNPs are null for association with SIgAD.

### 3.3 Meta-analysis of serum IgA data sets implicates IEI-related genes in IgA secretion

When additional studies are not available for meta-analysis, the use of methods which leverage information from GWAS of related traits can increase power. An obvious candidate for an informative auxiliary trait for SIgAD is serum IgA measured in the general population. To provide the most signal-rich serum IgA data set possible, we conducted the largest fixed-effect meta-analysis of serum IgA to date, combining two published studies [52,53] with our own GWAS of 7,938 subjects (Supplementary Figure 4 and Supplementary Table 5) to give a combined sample size of 57,063 (Supplementary Table 1). We identified 26 GWS associations of which eight were novel (Supplementary Figure 5). These included seven signals (five novel) which we linked to IEI genes (Tables S3 and S6): *FCGR3A/B*, *SANBR*, *IKZF1*, *POU2AF1*, *TRAF3*, *TNFSF13*, and *TNFRSF13B*.

As expected for an antibody-related phenotype, we were able to relate several novel associations to B-cell biology including a receptor-ligand pair. Lead SNP rs58647797 is located in an intron of *TNFRSF13B*, which encodes the transmembrane activator and calcium-modulator and cyclophilin ligand interactor (TACI), a receptor for B cell activating factor (BAFF) and A proliferation inducing ligand (APRIL) which induce T cell-independent class switch recombination [100,101] and plasma cell differentiation [102]. Complementary to this association was that of the SNP rs3803800, a missense variant in *TNFSF13* which encodes APRIL.

The lead SNP rs876038 is a *cis*-eQTL for *IKZF1* [91] lying in a promoter-interacting enhancer region upstream of the gene [103]. *IKZF1* encodes the Ikaros zinc finger 1 protein, a transcription factor and regulator of lymphocyte differentiation [104]. Ikaros is essential for pre-B cell differentiation [105] and immunoglobulin heavy chain rearrangement in mice [106]. We also identified an association with *IRF5*, a regulator of B-cell maturation and differentiation [107,108], and component of the IKZF1-IRF5 axis [109] which contributes to the antibody response [110]. IRF5 binds the *IKZF1* promoter to inhibit the gene’s expression [110].

Furthermore, we identified an association with the SNP rs9372120 in intron 6 of *ATG5*, which encodes the protein autophagy related 5 and regulates plasma cell differentiation [111]. The region hosting this SNP also participates in a looping interaction with *PRDM1* [112,113] which encodes the transcriptional repressor BLIMP-1, a regulator of plasma cell differentiation and antibody secretion [114–116]. We related another novel association with the intergenic SNP rs12713430 to *SANBR*, a negative regulator of class switch recombination [117], on the basis of its being a *cis*-eQTL for this downstream gene [91].

We could not confidently attribute the association of the SNP rs7522462 to a causal gene. The SNP is located in an intron of *INAVA*, which encodes the innate immune activator protein. This protein has roles in the regulation of epithelial cell junction stability at the mucosa [118,119] and pattern recognition signalling [120], and variants within it, including rs7522462, are associated with inflammatory bowel disease [121]. rs7522462 is also close to *GPR25*, which encodes an orphan G protein-coupled receptor expressed in natural killer and memory T cells, and is associated with several immune-mediated diseases [62,122,123].

Despite the serum IgA associations we could relate to IEIs, no significant association signals were shared between serum IgA and SIgAD, in contravention of our expectation of their sharing genetic effects (Table 1 and Supplementary Table 2). We did however find a significant negative genetic correlation between SIgAD and serum IgA, r_g_ = −0.28 (p = 0.009). The lack of overlap in significant associations likely reflects the limited power of our SIgAD GWAS; the genetic correlation provides a genome-wide estimate of pleiotropy which is not restricted to the effects of significantly associated loci.

### 3.4 SIgAD has significant genetic correlation with a range of immune-mediated diseases

A range of atopic and autoimmune diseases can be related to SIgAD on the basis of comorbidity, such as asthma, type 1 diabetes (T1D), rheumatoid arthritis, juvenile idiopathic arthritis, systemic lupus erythematosus (SLE), and inflammatory bowel disease (IBD) [124,125]. The sharing of common-variant genetic architecture among immune-mediated diseases (IMDs) is well-attested [126–128] and five of the nine loci we report to be associated with SIgAD in Table 1 are also associated with other IMDs (Supplementary Table 6).

We investigated whether this phenomenon was manifest genome-wide in SIgAD by estimating its genetic correlation with a panel of IMDs (Supplementary Table 1). In addition to using the genetic correlation to assess genetic similarity, we employed the nonparametric ‘genome-wide pairwise-association signal sharing’ (GPS) test [75,76]. We identified nine diseases with a significant positive genetic correlation with SIgAD whilst controlling the false discovery rate (FDR) at 0.05 (Supplementary Figure 6); all were also FDR-significant as assessed by the GPS test. These were rheumatoid arthritis, juvenile idiopathic arthritis, type 1 diabetes, Addison’s disease, primary biliary cholangitis, Crohn’s disease, ulcerative colitis, hypothyroidism, and IgA nephropathy.

IgA nephropathy (IgAN) is of interest as an IMD in which excess of serum IgA, rather than deficiency, is central to disease pathology [129]. In IgAN, deposition of galactose-deficient IgA1-containing immune complexes in the glomerular mesangium causes kidney injury [130]. IgAN showed a significant positive genetic correlation with both SIgAD (r_g_ = 0.30, p = 0.028) and serum IgA (r_g_ = 0.32, p = 2×10^-7^). Mendelian randomisation showed that IgAN’s genetic correlation with serum IgA represents a positive causal effect of serum IgA on the risk of IgAN (β = 0.94, p < 0.00034; Supplementary Figure 7); this causal effect has been anticipated in the literature by the association of higher serum IgA with IgAN [129]. While the estimated causal effect of serum IgA on SIgAD was negative (β = −0.29; Supplementary Figure 8), in concordance with the genetic correlation, it was not significant (p = 0.33). Despite the discordant directions of their genetic correlation with serum IgA and the estimated causal effect of IgA upon them, IgAN and SIgAD were positively correlated with one another (r_g_ = 0.30, p = 0.036), indicating a genetic component common to both which is not mediated by serum IgA levels.

### 3.5 Leveraging GWAS of related traits identifies 17 additional associations

We chose to condition the SIgAD meta-analysis on three traits which were correlated with SIgAD but relatively uncorrelated with one another, in order to capture different views of SIgAD’s genetic architecture and to satisfy the requirement of cFDR that one avoid conditioning on highly dependent traits. We complemented our SIgAD-IMD genetic correlation estimates by estimating the same parameter between IMDs to identify dependence between candidate auxiliary traits (Supplementary Figure 9).

We selected serum IgA, asthma, and rheumatoid arthritis as auxiliary traits on which to condition SIgAD (Supplementary Figure 6). SIgAD was positively correlated with asthma (r_g_ = 0.20), but this estimate was not nominally significant (p = 0.10). We chose asthma nonetheless for its status as the most prevalent atopy in SIgAD patients (19.1%) [18], its highly significant GPS test statistic (p = 0.0008), and our access to a highly-powered asthma GWAS for use as an auxiliary data set (Supplementary Table 1). Rheumatoid arthritis was highly correlated with SIgAD (r_g_ = 0.75) and is the third most common autoimmune manifestation in SIgAD patients (3.8%) after coeliac disease (6.6%) and inflammatory bowel disease (4.0%) [18]. The genetic correlation was low between all three conditioning traits (r_g_ ≤ 0.1).

We identified 17 additional associations using cFDR (Table 2 and Figure 3). Of these, five occurred in or near genes known to harbour IEI-causal variants: *CD247*, *IL2RA*, *IRF4*, *IKZF3*, and *PTPN2* (Supplementary Table 4).

**Figure 3.**
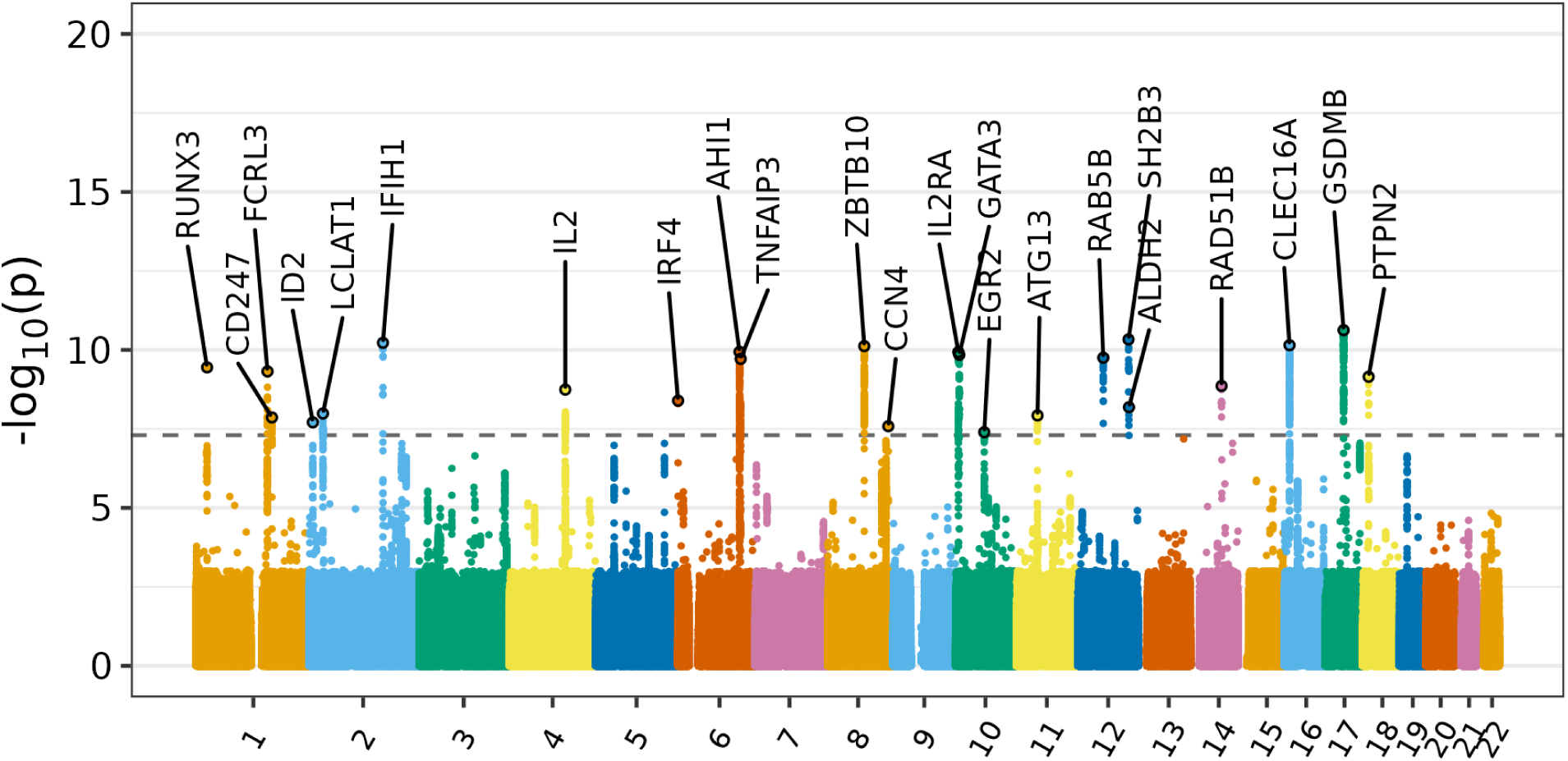
Manhattan plot depicting the results of the SIgAD cFDR analysis. Association signals are labelled with the genes to which they were mapped based on proximity and functional data.

**Table 2.**
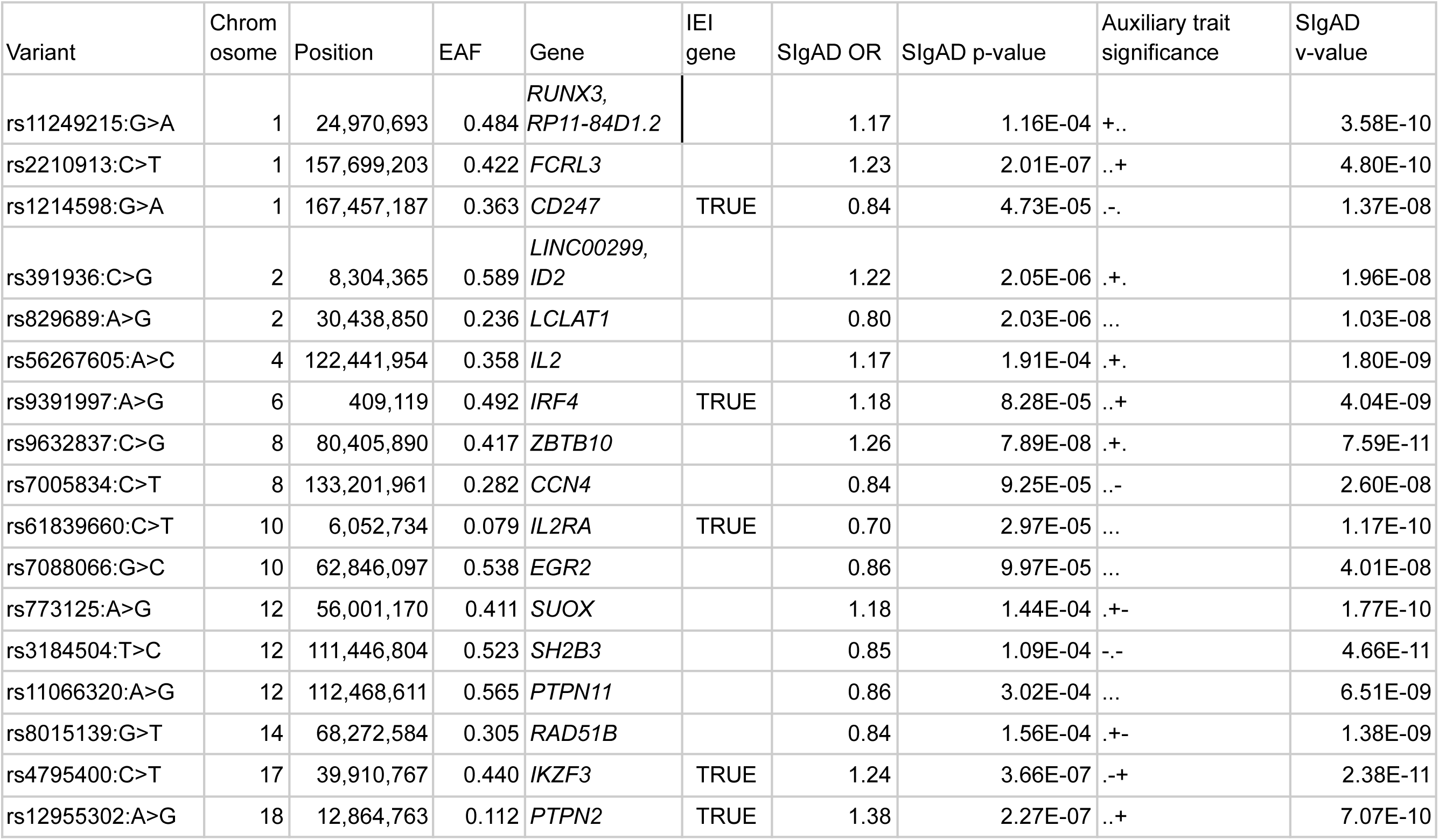
Lead SNPs from genome-wide significant associations in the cFDR analysis. The ‘Variant’ column gives the rsID of each SNP, and the reference and effect alleles separated by ‘>’. ‘EAF’ is effect allele frequency. Effect allele frequencies given were obtained from gnomAD’s estimate in non-Finnish Europeans. ‘Gene’ gives the gene(s) with the most evidence linking it/them to the association signal. ‘Novel’ indicates whether an association with SIgAD has previously been reported for a SNP. ‘IEI gene’ indicates whether the SNP is located in, near, or is otherwise associated with a gene known to harbour variants causal for IEIs. ‘OR’ is odds ratio. ‘Auxiliary trait significance’ indicates whether each auxiliary trait reached genome-wide significance for serum IgA, asthma, and rheumatoid arthritis, respectively: ‘+’ indicates a significant risk effect, ‘-’ a significant protective effect, and ‘.’ no significant effect. The ‘v-value’ is the output of the cFDR procedure and can be interpreted as a p-value against a null hypothesis of no association of the SNP with SIgAD after conditioning on the auxiliary traits.

The SNP rs1214598 is a *cis*-eQTL for *CD247* located in an intron of the same gene, which encodes the CD3ς chain of the T-cell receptor required for signal transduction. Loss-of-function mutations in *CD247* cause the severe combined immunodeficiency CD3ς deficiency [131]. The SIgAD signal in *CD247* appeared to have been modulated by the cFDR procedure due to our conditioning on asthma, with which this SNP is significantly associated [132]. rs1214598 is also in strong LD with SNPs associated with eczema [133] and autoimmune thyroid disease [134]. The SNP rs61839660 is located in intron 7 of *IL2RA* and is known for associations with multiple autoimmune [135] and atopic diseases including Crohn’s disease [136], type 1 diabetes [122], asthma and allergic disease [137], and eczema [138]. Loss-of-function mutations in *IL2RA* cause the autosomal recessive IEI CD25 deficiency, a disease of immune dysregulation featuring T regulatory cell defects, recurrent infection, lymphoproliferation, atopy, and autoimmunity [1,139,140].

rs9391997 is a 3’ UTR variant in *IRF4* associated with childhood-onset asthma [141], type 1 diabetes [62], autoimmune thyroid disease [134], and chronic lymphocytic leukaemia (CLL) [142]. It has been identified as one of several candidate causal variants in the *IRF4* 3’ UTR which may increase CLL risk through reduced *IRF4* expression [143]. *IRF4*, along with its homologue *IRF8*, plays a key role in the control of B-cell development [144]. *IRF4* is important in plasma cell differentiation and class switch recombination, acting as a crucial ‘transcriptional switch’ in the development of plasma cells [145]; SIgAD and CVID may both see failure of these processes. IRF4 haploinsufficiency is an autosomal dominant IEI and defect of innate immunity affecting leukocytes and macrophages which confers susceptibility to the rare bacterial infection Whipple’s disease [1,146].

Our cFDR analysis also elevated to genome-wide significance the SNP rs4795400, located in an intron of *GSDMB* and strongly associated with childhood-onset asthma [137,147]. However, the lead SNP in this association signal in the SIgAD meta-analysis, rs34073687 (p = 1.51e-7), occurred in an intron of *IKZF3*. This gene plays a role in B-cell development [148] and has recently been reported to harbour missense variants causing an autosomal dominant IEI, the combined immunodeficiency AIOLOS deficiency [1,149,150]. This IEI features impaired differentiation of B cells, B-cell lymphopaenia and malignancy, hypogammaglobulinaemia, and recurrent infections with susceptibility to Epstein-Barr virus, and the opportunistic pathogens *Pneumocystis jirovecii* and *Mycobacterium avium* complex [151].

Another SNP, rs12955302, was close to significance in the SIgAD meta-analysis (p = 2.3e-7) before reaching it in the cFDR analysis. rs12955302 is located in an intron of *PTPN2*, an immunologically salient gene, and is associated with a range of IMDs including Crohn’s disease, ulcerative colitis [121], T1D [62], and coeliac disease [152]. This SNP is in LD with others associated with rheumatoid arthritis [60]. *PTPN2* encodes protein tyrosine phosphatase 2, which negatively regulates cytokine and T-cell receptor signalling and thereby restrains T cell activation [153]. *Ptpn2*-deficient mice display systemic inflammation and autoimmunity [154] whilst reduced *Ptpn2* expression can exacerbate autoimmunity [155]. A single case of CVID has been attributed to the combination of a rare stop-gain and common hypomorphic variant in *trans* in *PTPN2*; the stop-gain mutation was seen in the patient’s mother, in whom it appeared to cause a complex autoimmune phenotype featuring insulin-dependent diabetes, SLE, hypothyroidism, and neutropaenia, whilst the hypomorphic variant appeared in the patient’s brother, who suffered from severe allergic nasal polyposis [156].

In addition to those associations we could relate to monogenic IEIs, the cFDR analysis identified novel associations shared with IgAN and serum IgA. The sub-threshold SIgAD signal (p = 2e-7) in *FCRL3* shared with IgAN was elevated to genome-wide significance in the cFDR analysis. *FCRL3* encodes Fc receptor-like protein 3 (FCRL3), which is preferentially expressed in B cells and both inhibits B-cell receptor signalling [157] and mediates B cell activation in response to innate immune signals [158]. Secretory IgA was recently identified as an FCRL3 ligand: when expressed on T regulatory cells, FCRL3’s binding secretory IgA induces a pro-inflammatory phenotype and may thereby stimulate immune responses to the loss of mucosal integrity [159]. Serum IgA production has a strong, reproducible association with *RUNX3* and our cFDR analysis elevated to significance an association of SIgAD with the same locus: the sole genome-wide significant SNP lay in the exonic sequence of the obscure lncRNA RP11-84D1.2 and within a candidate cis-regulatory element. Transforming growth factor Β-mediated IgA class switching is dependent on *RUNX3* [160], whilst Runx3-deficient mice develop IBD and produce increased quantities of IgA [161].

The SNP rs9632837 was near significance in our SIgAD meta-analysis (p = 7.9e-8) and became significant in the cFDR analysis. This SNP lies in an lncRNA and is an eQTL for *ZBTB10* [91], which encodes a zinc finger protein. It has been argued that ZBTB10’s repression of the Sp1 transcription factor, which regulates a number of cytokine-related genes, may account for the association of this gene with asthma [162]. Other members of the ZBTB family transcription factors are known to regulate B-cell development [163].

## 4. Discussion

We have investigated the common-variant architecture of SIgAD, an IMD which is comparatively common in many ancestries but rare with respect to its representation in IEI cohorts. As of 2018, the UK PID Registry featured 85 SIgAD patients among 4,242 cases listed by diagnosis (prevalence 1:300 to 1:1,200 [5]); the second most common IEI, CVID (prevalence 1:25,000 [164]), accounted for 1,273 patients [165]. Meaningful increases in the sample size of SIgAD GWAS in the immediate future thus seem unlikely. The small increase in sample size in our meta-analysis allowed us to identify four novel associations, but three of these were only marginally below the threshold of genome-wide significance in the GWAS by Bronson and colleagues. The fourth association we report, mapped to *TNFAIP3*, was significant but not identified in their work, possibly owing to its proximity to the *AHI1* signal. We believe further incremental increases in sample size would be unlikely to yield as many novel associations.

The multiplicity of loci identified in our meta-analysis and cFDR analysis strengthen the evidence for a polygenic, common-variant aetiology for SIgAD. *CD86* is of note as the first SIgAD gene identified by GWAS to relate to co-stimulation and T-/B-cell interaction in the disease. Impaired upregulation of CD86 on naive B cells in response to stimulation has been reported in a subset of CVID patients [166,167], a disease thought to be genetically related to SIgAD on the basis of the co-occurence of both disorders in the same families [3,33,36,168,169], the similarity in underlying immune dysfunction, the progression of SIgAD to CVID [170], and evidence of shared genetic associations [41,168]. Disruption of T-/B-cell interactions in the germinal centre through deficient CD86-mediated costimulation might inhibit plasma cell differentiation and class switching, processes thought to be dysfunctional in SIgAD. However, a study of B cells in three SIgAD patients found no difference in the upregulation of CD86 in response to antigenic stimuli compared to healthy controls [17].

T-cell biology was further implicated by our identification through cFDR of SIgAD-associated SNPs in *PTPN2*, *CD247*, and the receptor subunit and ligand pair *IL2RA* and *IL2*. With the exception of *IL2*, these genes have known associations with T cell-mediated autoimmunity and Mendelian IEIs. Their identification may provide a partial genetic explanation for the elevated burden of autoimmunity in SIgAD and the presence of anti-IgA antibodies in ∼37% of cases [171]. More in keeping with the predominant, B cell-focused understanding of SIgAD was our discovery of associated SNPs in *IKZF3*, *IRF4*, and *RUNX3*, genes with key roles in B-cell development, plasma cell differentiation, and class switching. Both *IKZF3* and *IRF4* also harbour variants causal for Mendelian IEIs.

The strength of SIgAD’s genetic correlation with IMDs such as type 1 diabetes and rheumatoid arthritis suggests these diseases share latent common-variant effects with SIgAD which our meta-analysis was insufficiently powered to detect. Detailed exploration of shared loci can elucidate shared pathways and mechanisms of disease in a way that estimation of the genetic correlation, a single genome-wide parameter, cannot. Given the positive correlation between SIgAD and IgAN, the opposing signs of their genetic correlation with serum IgA were notable. They suggest that there exist latent IMD-related effects which are distinct from those genetic effects which act through serum IgA.

We report novel associations of serum IgA with SNPs in *TNFRSF13B* and *TNFSF13*, which encode the members of the receptor-ligand pair TACI and APRIL, respectively. Both of these genes have been the subject of investigation in SIgAD: a role for missense TACI variants in SIgAD has been proposed and disputed in the literature [41,45] whilst the APRIL SNP rs3803800 has previously tested negative for association with SIgAD in two small cohorts of 164 and 51 patients [172]. TACI mediates T cell-independent class switching [100,101] and plasma cell differentiation [102], both processes identified as dysfunctional in SIgAD. IgA class switching is also impaired in APRIL-deficient mice [173] and the sole known human case of APRIL deficiency presented with low IgA as part of a CVID phenotype [174]. Neither *TNFRSF13B* nor *TNFSF13* harboured significantly associated SNPs in our SIgAD analyses, but the association of SNPs in both with serum IgA, the genetic correlation between serum IgA and SIgAD, and the functions of this signalling pathway suggest larger GWAS might find SIgAD-associated variants in these genes.

Serum IgA measured in the general population represents an ostensibly informative trait for analysis with SIgAD, but the most salient clinical manifestation of SIgAD, susceptibility to infection, is a consequence of deficiency in secretory IgA. Serum and secretory IgA status may be discordant: faecal IgA has been detected in the absence of serum IgA in a small 19-subject paediatric study and faecal IgA status in the same patients better correlated with disease [175]. The majority of serum IgA-deficient individuals are asymptomatic [176]. Severity of disease in SIgAD appears related to the presence of a concomitant IgG subclass deficiency in a notable minority of cases [177]. Whilst we successfully leveraged serum IgA data to explore SIgAD in this work, our understanding of SIgAD would likely further benefit from GWAS of secretory IgA levels and serum IgG subclass levels, data sets which are as yet publicly unavailable. Our cFDR analysis pointed to the importance of secretory IgA in its identification of *FCRL3,* recently shown to encode a secretory IgA receptor.

Our identification of additional common-variant SIgAD associations will complement the search for rarer variants insofar as it expands the list of candidate loci to be examined for their presence. We have shown IEI genes are enriched for common variants indicating evidence of association with SIgAD. We hypothesise the converse: genes harbouring common SIgAD-associated variants, such as *CLEC16A*, *FAS*, and *IFIH1*, might prove natural candidates for the identification of rarer, more penetrant variants associated with SIgAD, and IEIs more generally. *IFIH1* in particular is already known to harbour both a common SIgAD-associated variant and, at least in the case of one patient, a *de novo* gain-of-function missense mutation leading to systemic lupus erythematosus and IgA deficiency [178].

Unlike in many IEIs, highly penetrant mutations causing SIgAD have not been identified. Absence of evidence for monogenic explanations is not evidence of their absence and IgA deficiency occurs as part of other more complex monogenic conditions, such as ataxia telangiectasia. Nonetheless, the failure of decades of study to definitively identify such genes for SIgAD outside of the MHC does motivate further efforts at common variant discovery. A number of monogenic defects have been identified in CVID, but a complex, polygenic aetiology is still suggested for the sporadic cases which make up around three-quarters of patients [179].

Given the possibility SIgAD to CVID progression, the incidence of IgA deficiency in many CVID patients and of IgG subclass deficiency in some SIgAD cases, and the known sharing of loci such as *CLEC16A* [180], the understanding of both diseases could benefit from an analysis of their shared genetic architecture. A comprehensive analysis would be subject to the availability of more highly-powered studies: published GWAS of CVID have sample sizes even smaller than those of SIgAD [180,181]. As we have shown in the present work, the cFDR procedure represents a useful tool to improve power in the absence of larger rare disease cohorts.

## 5. Conclusions

We have expanded the number of known SIgAD-associated loci and in doing so bolstered the case for a common-variant genetic aetiology for the disease. We have documented evidence of SIgAD’s shared genetic architecture with serum IgA production, IMDs, and a number of monogenic inborn errors of immunity, illustrating the significance of variant and gene pleiotropy in the architecture of immune-related traits. Our successful application of the cFDR procedure to the problem of insufficient power shows how variant pleiotropy can be exploited to overcome this small sample sizes, whilst the enrichment of SIgAD association signal in IEI genes suggests the exploration of these for the identification of as-yet elusive rare variants associated with SIgAD.

## Data Availability

GWAS data sets, cFDR results, GPS test statistics, and genetic correlation estimates are available at the following Zenodo page: https://zenodo.org/doi/10.5281/zenodo.11929771.

## Acknowledgements

We would like to thank our colleague Dr Guillermo Reales for his creation of the *GWAS_tools* pipeline. We also thank Christopher Benson, Debora Lucarelli, and Jing Hua Zhao for their contribution to the generation of data used in this paper. We wish to acknowledge all GWAS participants, in particular those of the UK Biobank and FinnGen, for their contribution to the data used herein. We also acknowledge the investigators who carried out these GWAS and made their summary statistics publicly available. We acknowledge in particular the Pan-UKBB team [54]. We are grateful to all the participants of the EPIC-Norfolk study and to the many members of the study teams at the University of Cambridge who have enabled this research.

## Funding

This work was supported by the Wellcome Trust (WT107881) and the Medical Research Council (MC_UU_00002/4, MC_UU_00040/01).

The EPIC-Norfolk study (DOI 10.22025/2019.10.105.00004) has received funding from the Medical Research Council (MR/N003284/1, MC-UU_12015/1, and MC_UU_00006/1) and Cancer Research UK (C864/A14136). The genetics work in the EPIC-Norfolk study was funded by the Medical Research Council (MC_PC_13048). We also acknowledge funding from the European Union’s Horizon 2020 Research and Innovation Programme under grant agreement 633964 (ImmunoAgeing).

For the purpose of Open Access, the author has applied a CC BY public copyright licence to any Author Accepted Manuscript version arising from this submission.

## Conflicts of Interest

C.W. receives funding from GSK and MSD and is a part time employee of GSK. These companies had no input into this work.

## Supporting information

### Supplementary tables

**Supplementary Table 1.** The GWAS data sets included in the SIgAD and IgA meta-analyses, and SIgAD cFDR analysis. Pan-UKB is the Pan-UK Biobank study. IMSGC is the International Multiple Sclerosis Genetics Consortium.

**Supplementary Table 2.** Lead SNPs from genome-wide significant associations in the serum IgA GWAS meta-analysis. The ‘Variant’ column gives the rsID of each SNP, and the reference and effect alleles separated by ‘>’. Effect allele frequencies given were obtained from gnomAD’s estimate in non-Finnish Europeans. ‘Gene’ gives the gene(s) with the most evidence linking it/them to the association signal. ‘Novel’ indicates whether an association with SIgAD has previously been reported for a SNP. ‘IEI gene’ indicates whether the SNP is located in, near, or is otherwise associated with a gene known to harbour variants causal for IEIs. ‘GWAS p-value’ gives the meta-analytic p-value. ‘Effect direction’ indicates whether the effect allele is associated with an IgA-increasing (’+’) or decreasing (’-’) effect. ‘Study effects’ indicates whether a significant effect was found in the component GWAS of our meta-analysis: Liu, our own GWAS, and Dennis, respectively. ‘.’ indicates no significant effect.

**Supplementary Table 3.** The ‘Variant’ column gives the rsID of each SNP, and the reference and effect alleles separated by ‘>’. ‘Effect size’ gives the GWAS effect estimate for the SNP on the parenthetically named phenotype and ‘Standard error’ gives the corresponding standard error of the effect estimate. ‘IgA’ is serum IgA and ‘IgAN’ is IgA nephropathy.

**Supplementary Table 4.** ‘Gene’ gives the name of the IEI-associated gene. ‘Analysis’ gives the phenotype with which the gene was associated and the study modality (GWAS meta-analysis or cFDR) used to identify the association in this work. ‘Inborn error of immunity’ gives the IEI with which the gene is associated. ‘IgA deficiency’ indicates whether IgA deficiency is known to be a feature of the IEI.

**Supplementary Table 5.** Lead SNPs from genome-wide significant associations in our GWAS of serum IgA. The ‘Variant’ column gives the rsID of each SNP, and the reference and effect alleles separated by ‘>’. Effect allele frequencies given were obtained from gnomAD’s estimate in non-Finnish Europeans. ‘Gene’ gives the gene(s) with the most evidence linking it/them to the association signal. ‘Novel’ indicates whether an association with serum IgA had previously been reported for a SNP. ‘Effect direction’ indicates whether the effect allele is associated with an IgA-increasing (’+’) or decreasing (’-’) effect.

**Supplementary Table 6.** The association of the lead SNPs from the SIgAD meta-analysis with other immune-mediated diseases (IMDs) as identified by GWAS. The ‘Variant’ column gives the rsID of each SNP, and the reference and effect alleles separated by ‘>’. ‘Gene’ gives the gene(s) with the most evidence linking it/them to the association signal. ‘Novel’ indicates whether an association with SIgAD has previously been reported for a SNP. ‘IMD associations in LD’ lists IMDs which have significantly associated variants in linkage disequilibrium with the SIgAD-associated variant given in ‘Variant’.

**Supplementary Table 7.** ‘Catalog summary’ denotes the summary table of lead SNPs provided on the EBI GWAS Catalog. ‘Bronson’ indicates the GWAS summary statistics downloaded from the EBI GWAS Catalog. ‘Lim’ indicates the GWAS summary statistics we obtained by analysing the genotype data published by Lim et al. ‘T2’ and ‘ST2’ denote Table 2 and Supplementary Table 5 of Bronson et al. ‘OR’ denotes odds ratio. Odds ratios ‘OR A’ and ‘OR B’ were computed from allele counts in Supplementary Table 5 of Bronson et al. taking the ‘A’ and ‘B’ labels as the effect allele, respectively.

### Supplementary figures

**Supplementary Figure 1.**
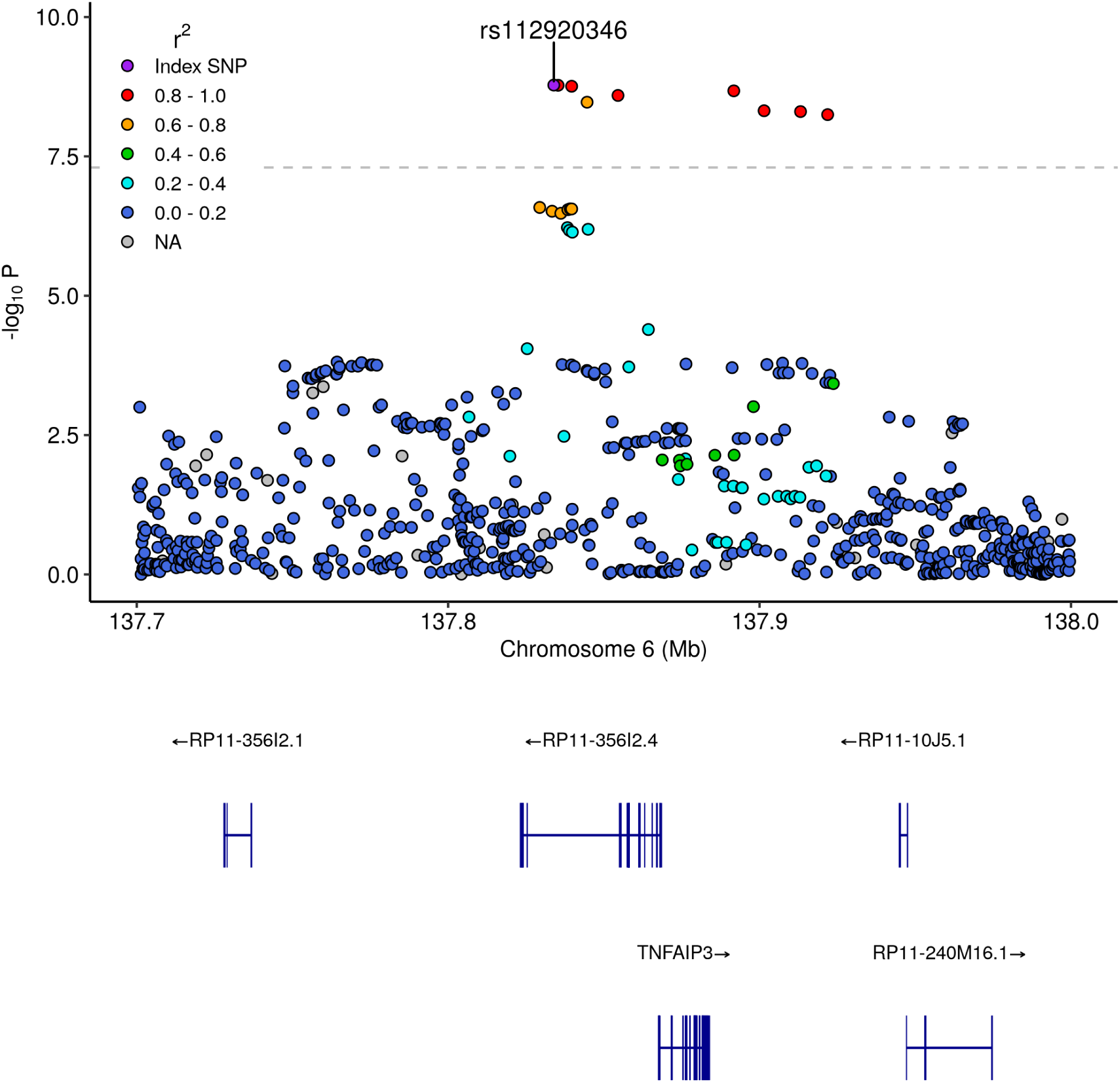
The *TNFAIP3* association signal. Each point corresponds to a SNP’s p-value from a test of association in the GWAS meta-analysis. The points are coloured according to their squared correlation (r^2^) with the lead SNP rs112920346. The *WAKMAR2* gene is RP11-356I2.4.

**Supplementary Figure 2.**
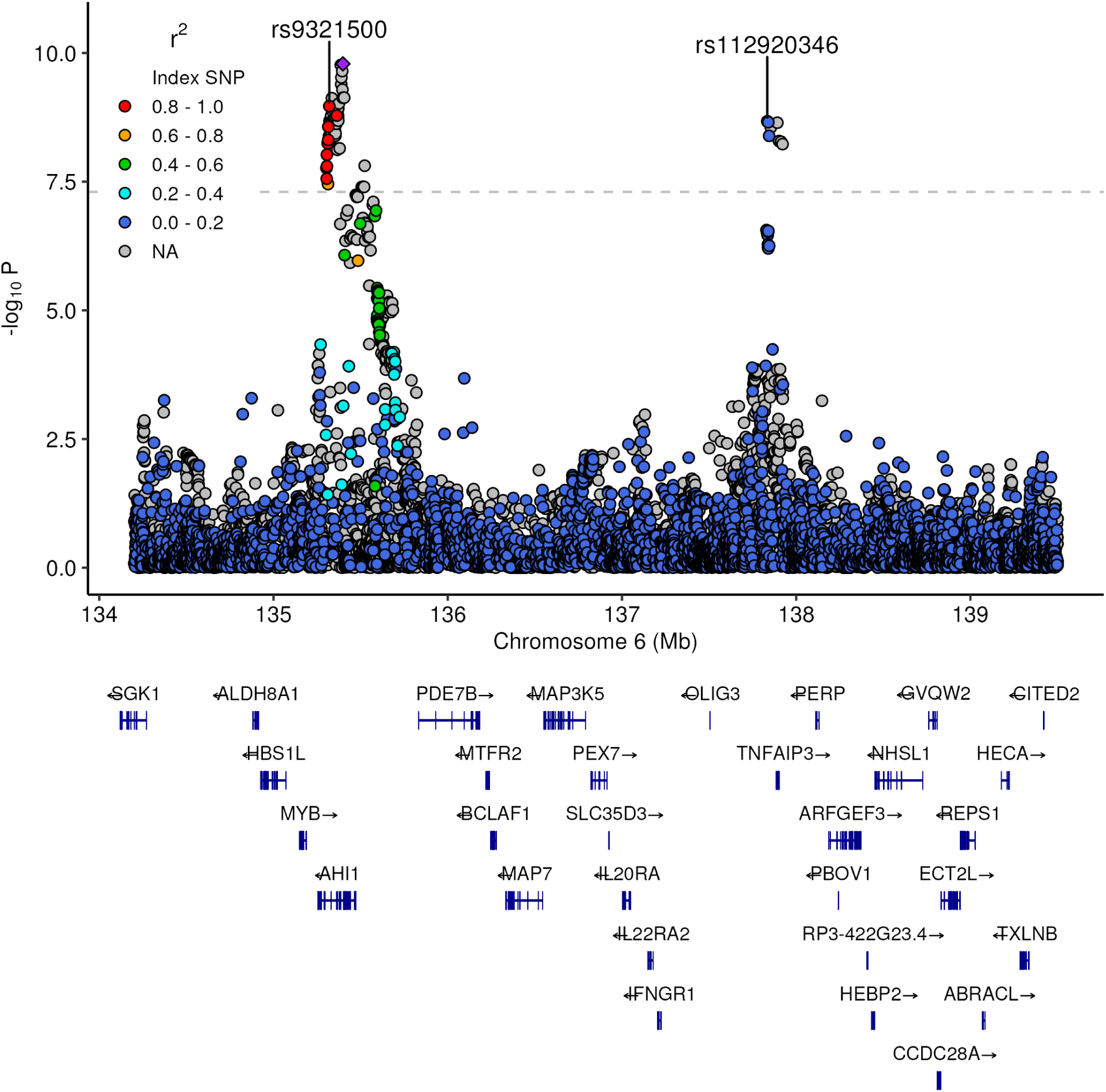
The *TNFAIP3* signal was distinct from that which was previously reported at *AHI1*. Each point corresponds to a SNP’s p-value from a test of association in the GWAS meta-analysis. The points are coloured according to their squared correlation (r^2^) with rs9321500, the SNP in the *AHI1* signal for which LD information was available and which had the smallest p-value. SNPs for which LD information was not available are coloured grey. Only protein-coding genes are depicted.

**Supplementary Figure 3.**
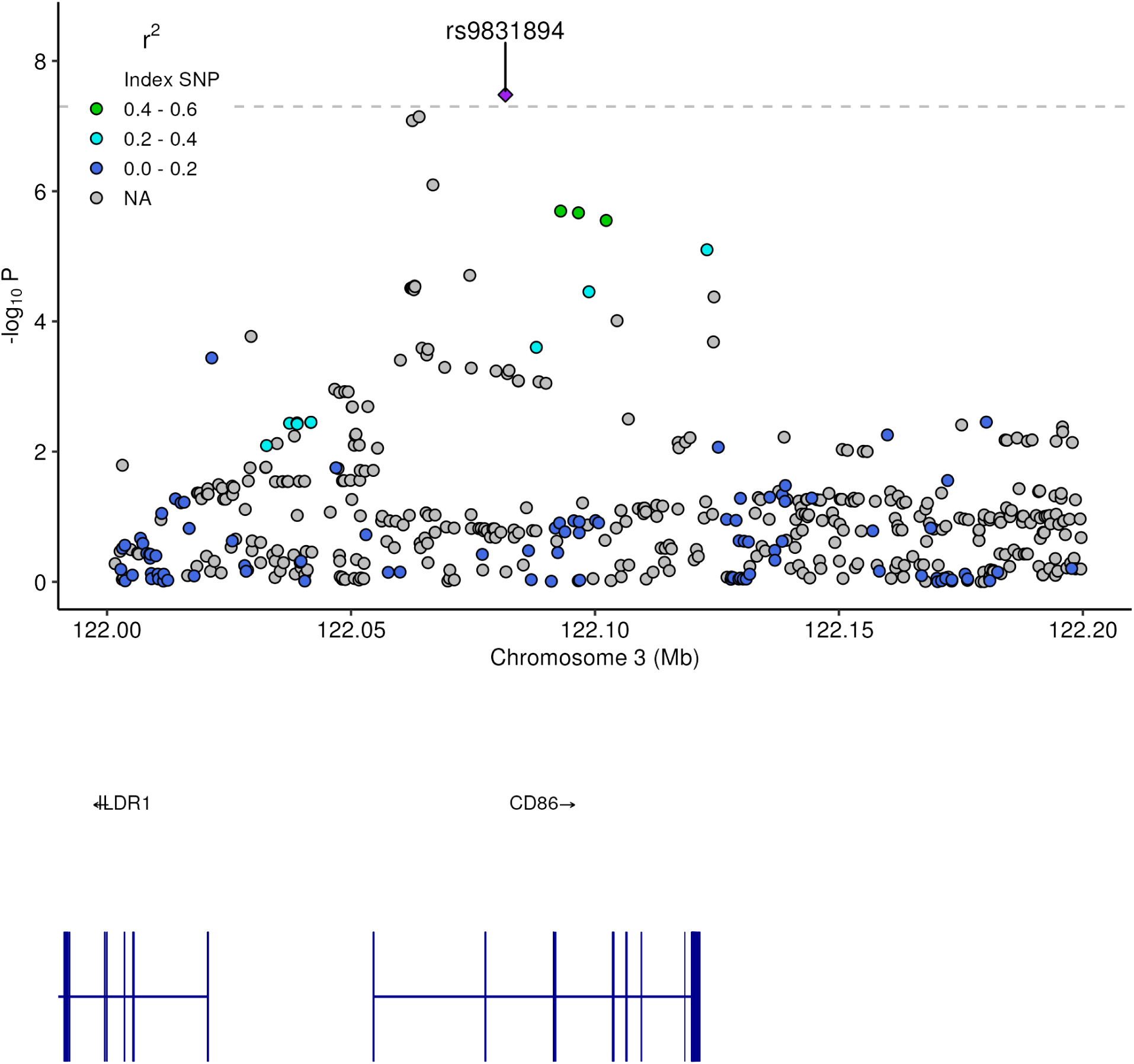
The *CD86* GWAS association signal. Each point corresponds to a SNP’s p-value from a test of association in the GWAS meta-analysis. The points are coloured according to their squared correlation (r^2^) with the lead SNP rs983189.

**Supplementary Figure 4.**
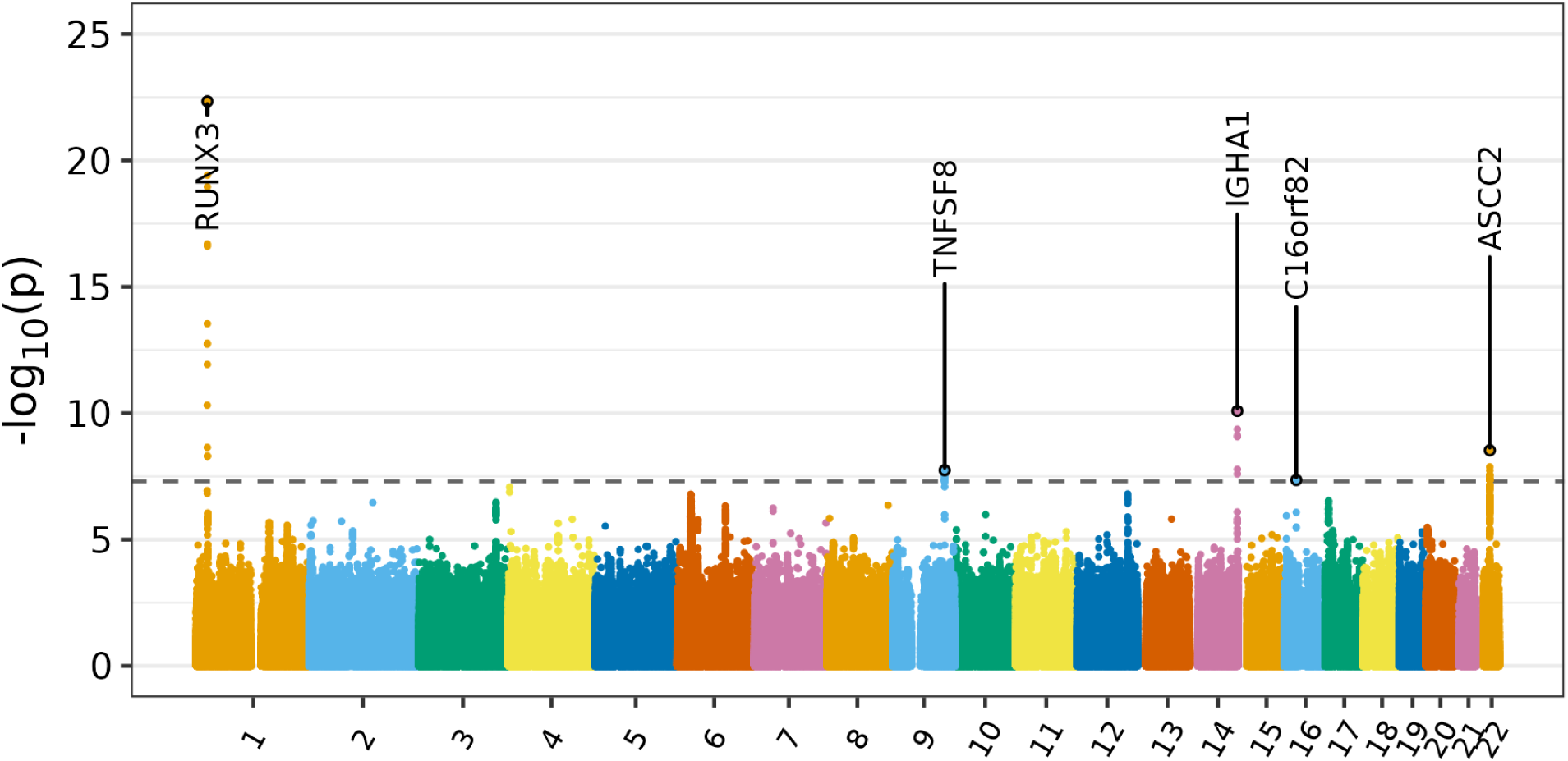
Manhattan plot depicting the results of our GWAS of serum IgA. Association signals are labelled with the genes to which they were mapped based on proximity and functional data.

**Supplementary Figure 5.**
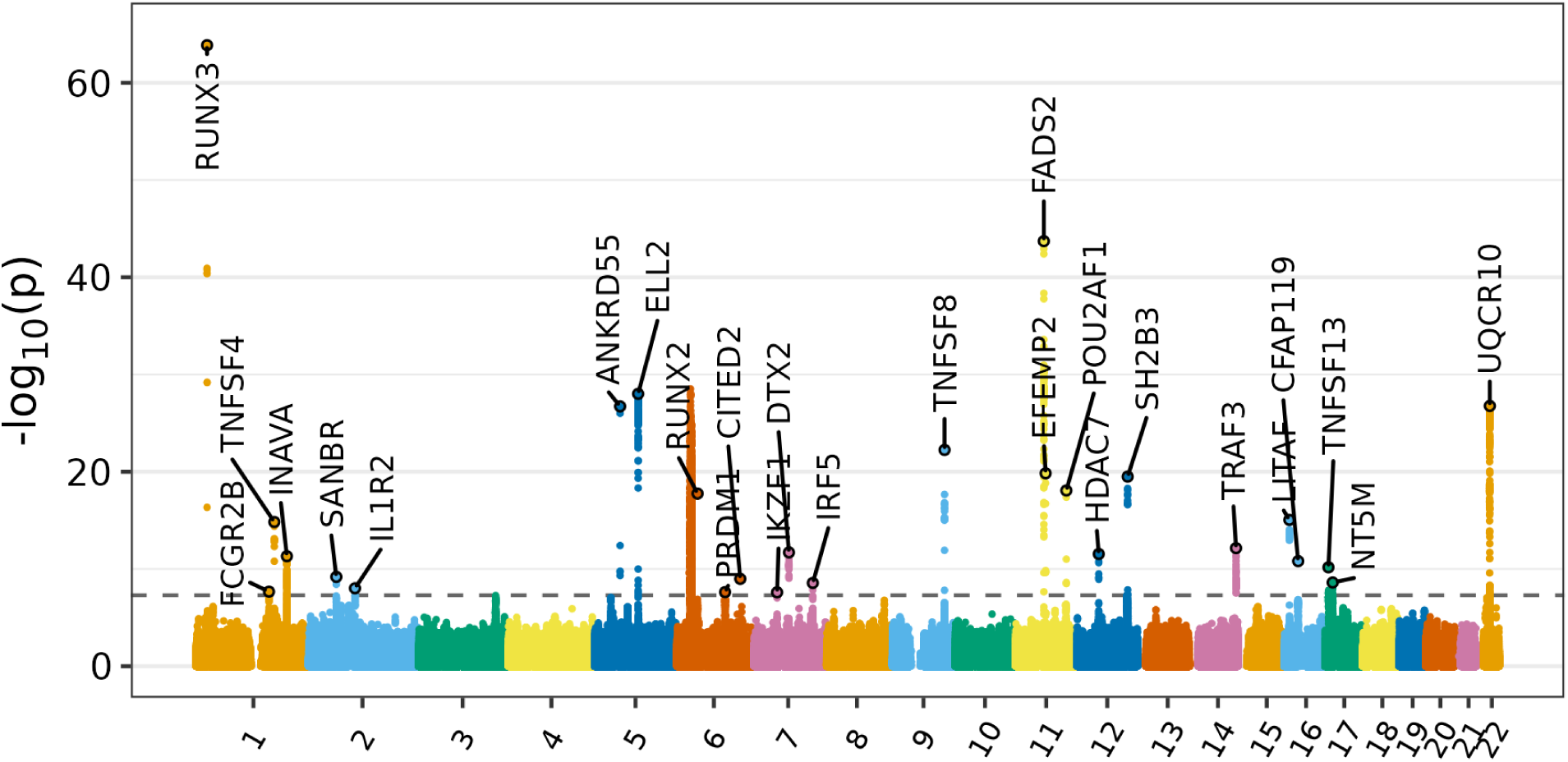
Manhattan plot depicting the results of the GWAS meta-analysis of serum IgA. Association signals are labelled with the genes to which they were mapped based on proximity and functional data.

**Supplementary Figure 6.**
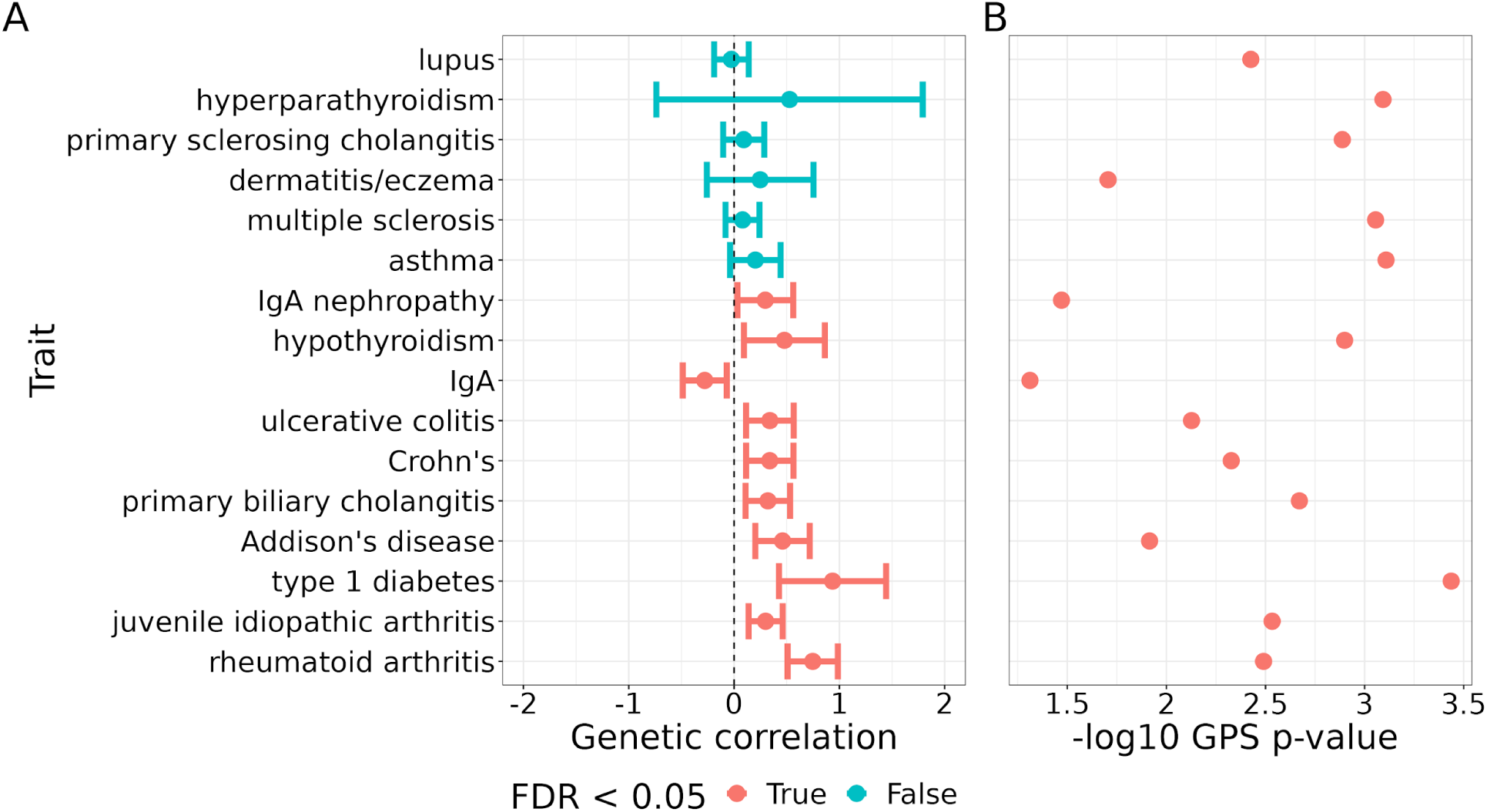
Estimates of SIgAD’s genetic correlation with a panel of immune traits and p-values from application of the GPS test to SIgAD and members of the same panel. ‘A’ depicts the genetic correlation estimates with 95% confidence intervals. Traits are ordered by the (uncorrected) p-value of a test of non-zero genetic correlation with SIgAD. The colour of the point indicates whether the null was rejected for a test of non-zero genetic correlation with SIgAD (A) or the GPS test (B) whilst controlling FDR at 0.05.

**Supplementary Figure 7.**
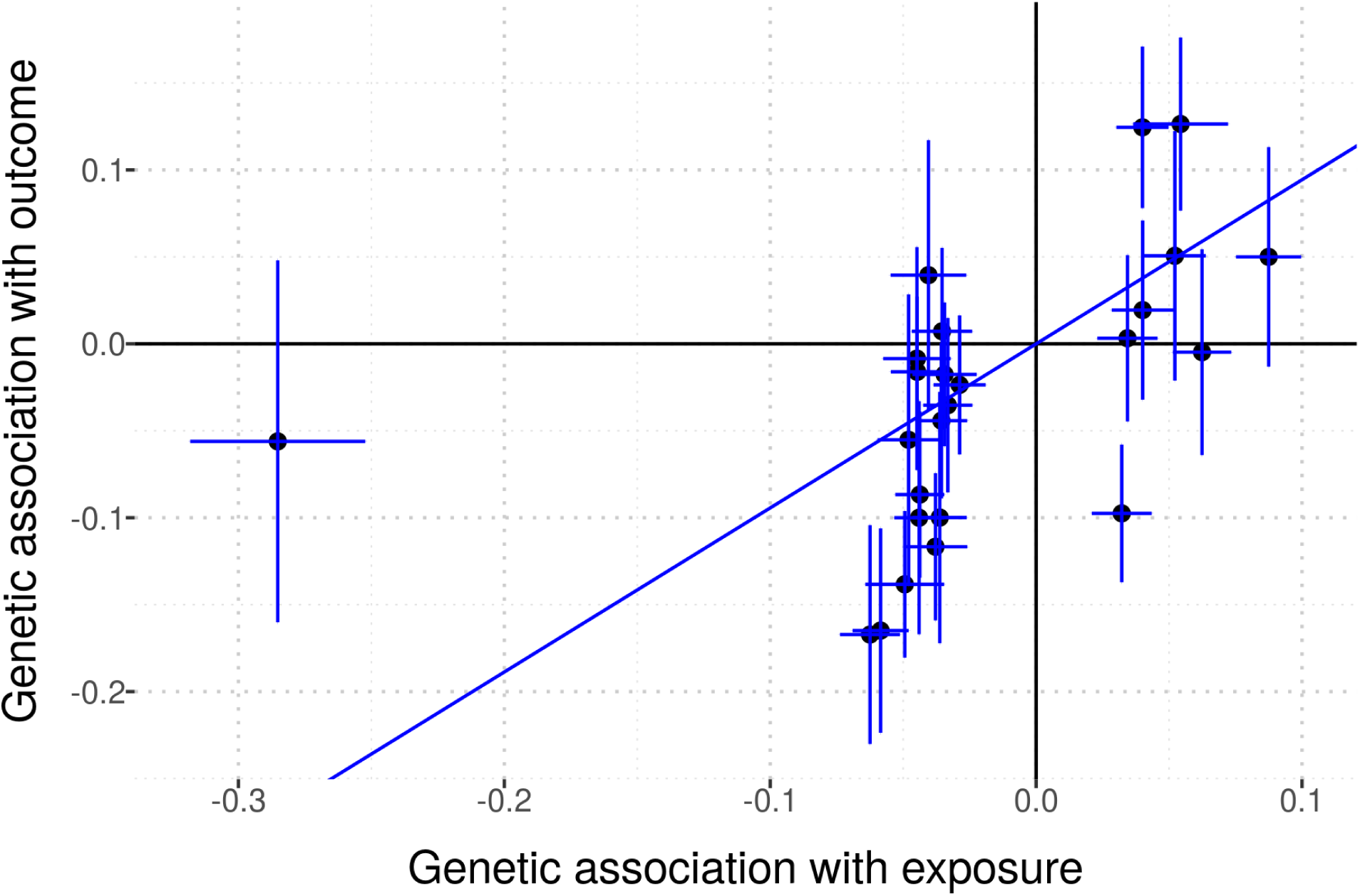
Mendelian randomisation of serum IgA (the exposure) and IgA nephropathy (the outcome). Each point corresponds to a SNP which was significantly associated with serum IgA in our meta-analysis. The x-coordinate is given by the effect estimate of the SNP for serum IgA and the y-coordinate by its effect estimate for IgA nephropathy. The blue lines give 95% confidence intervals for the effect estimates for each phenotype. The gradient of the line passing through the origin gives the estimate of the causal effect of serum IgA on the risk of IgA nephropathy, 0.94 (p < 0.00034).

**Supplementary Figure 8.**
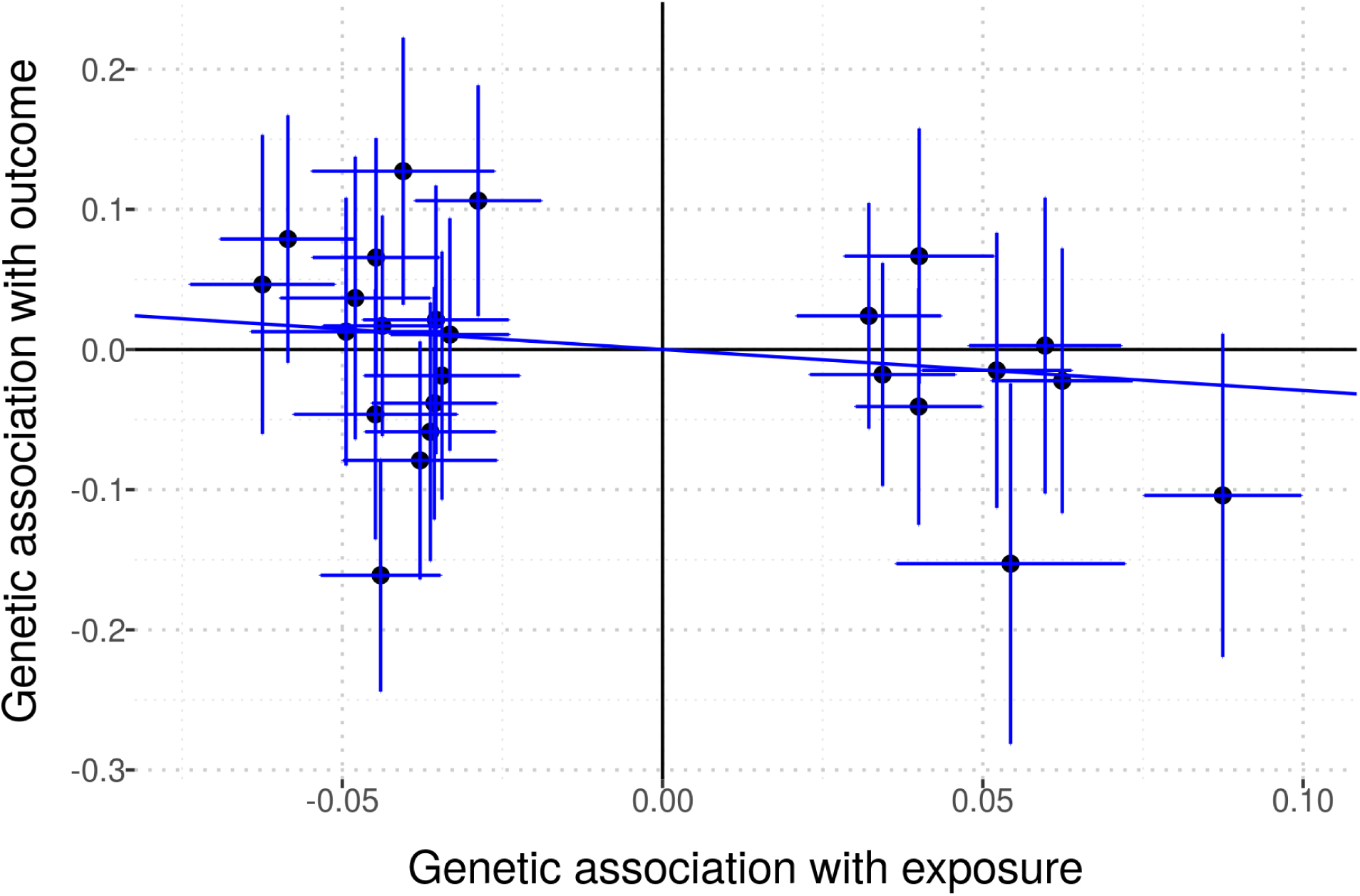
Mendelian randomisation of serum IgA (the exposure) and SIgAD (the outcome). Each point corresponds to a SNP which was significantly associated with serum IgA in our meta-analysis. The x-coordinate is given by the effect estimate of the SNP for serum IgA and the y-coordinate by its effect estimate for SIgAD. The blue lines give 95% confidence intervals for the effect estimates for each phenotype. The gradient of the line passing through the origin gives the estimate of the causal effect of serum IgA on the risk of SIgAD, −0.29, which was not significant (p = 0.33).

**Supplementary Figure 9.**
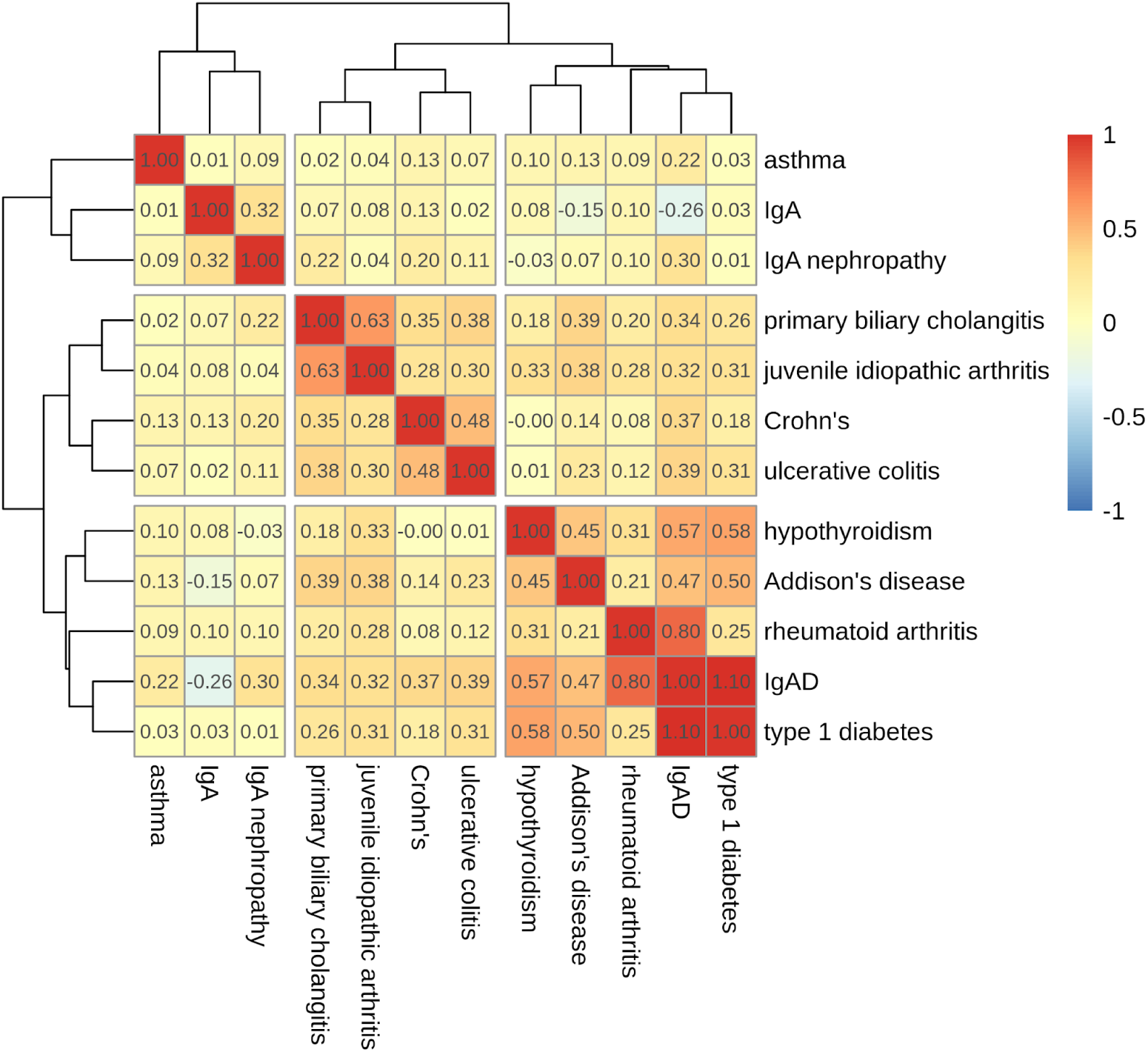
Genetic correlation estimates among SIgAD (‘IgAD’) and selected immune traits. The traits were hierarchically clustered into three groups on the basis of our choice of three conditioning traits for the cFDR analysis.

**Supplementary Figure 10.**
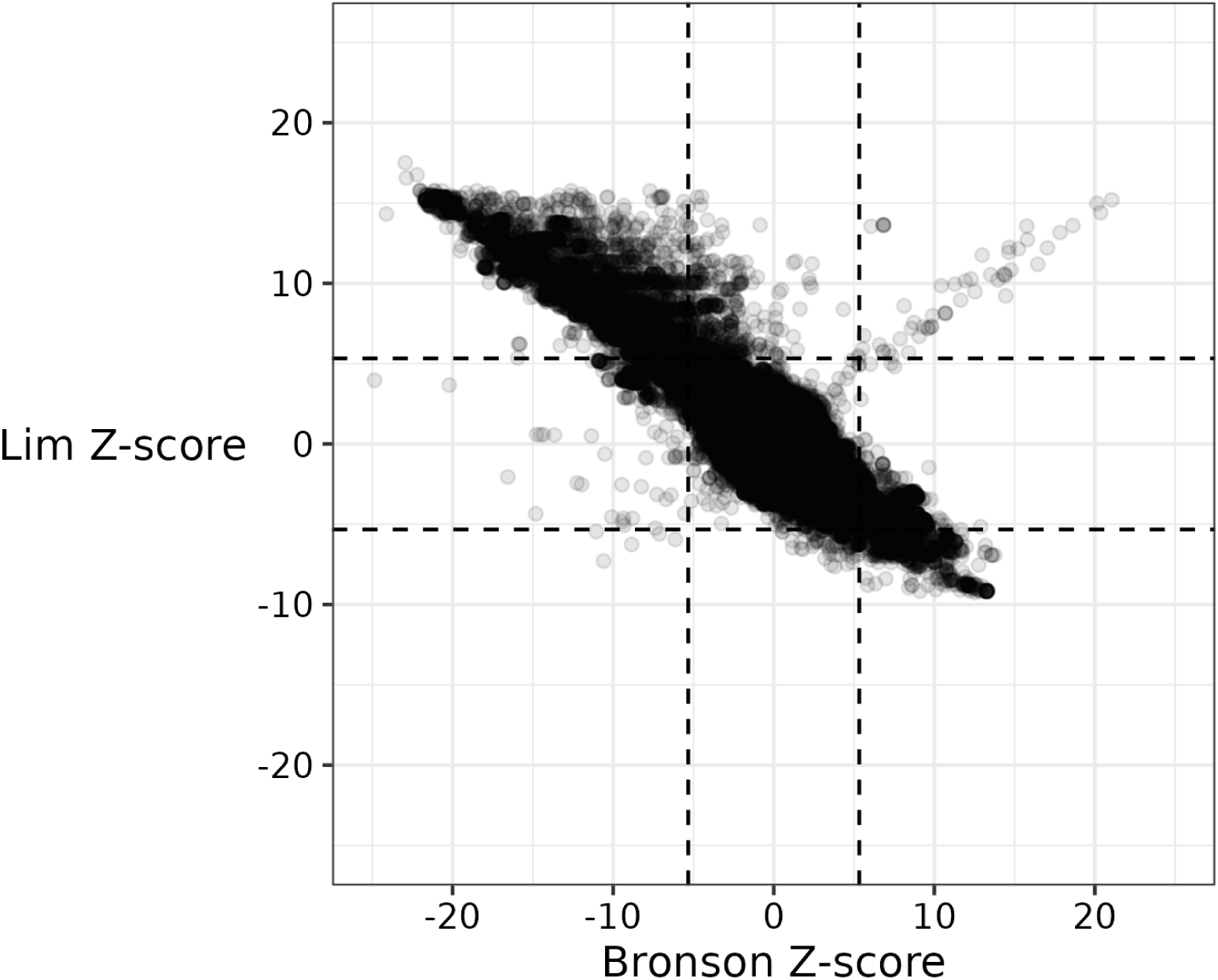
Each point corresponds to a SNP common to both Bronson and colleagues’ SIgAD GWAS and our GWAS of the SIgAD genotype data published by Lim and colleagues. The x- and y-coordinate of each point are given by the Z-score of the SNP in the Bronson and Lim GWAS, respectively.

### Supplementary data

**Supplementary Data 1.** IDs, names, and genomic coordinates of the 448 IEI genes used in enrichment analysis. *id* is the Ensembl gene ID. *tss* gives the position of the transcription start site. *fwdStrand* indicates whether the gene is found on the forward or reverse strand.

### Supplementary methods

#### Detection and correction of mislabelled columns in the Bronson data set

We observed an apparent mislabelling of the two allele columns in the version of Bronson and colleagues’ SIgAD GWAS data set available on the GWAS Catalog. This mislabelling reversed the labels of the ‘effect’ and ‘other’ allele columns without also taking the reciprocal of the odds ratio, with the consequence that the odds ratio estimate reported at each SNP was the multiplicative inverse of the true odds ratio estimate expected for the given alleles. We determined these columns were mislabelled after cross-referencing several sources of information relating to four non-MHC lead SNPs reported: the summary statistics themselves, the GWAS Catalog’s summary of lead SNPs, and Table 2 and Supplementary Table 5 of Bronson and colleagues’ paper (Supplementary Table 7). The Catalog summary reported the same odds ratio estimates as those in the summary statistics, but reported as risk (i.e. ‘effect’) alleles those alleles labelled ‘other’ alleles in the summary statistics.

In Supplementary Table 5 of their paper, Bronson et al. provided allele frequencies and odds ratios for these lead SNPs in each national subset making up the study cohort. The alleles were labelled only as ‘A’ and ‘B’, and the direction of the odds ratio with respect to these was not specified. We recomputed the allelic odds ratios using the information given in Supplementary Table 5 for the Swedish cohort (the largest national cohort) and used the resulting ratios to disambiguate the direction of the stated odds ratios. These transpired to have as their effect allele whichever was the minor allele in cases, such that neither ‘A’ nor ‘B’ was consistently designated the effect allele for the stated odds ratios. We found that odds ratios with allele ‘B’ as the effect allele matched the direction of effect for the odds ratios reported in the Catalog summary and the summary statistics, and that allele ‘B’ matched the Catalog summary’s reported risk allele but the summary statistics’ ‘other’ allele. These findings (restricted as they are to only four variants) are consistent with the summary statistics’ allele column labels having been reversed without a concomitant modification of the corresponding odds ratios.

We corroborated these findings by performing a GWAS of SIgAD using genotype data published by Lim and colleagues [182] on the European Nucleotide Archive under project accession PRJEB4929. The cases in this cohort appear to correspond to a subset of the cohort studied by Bronson and colleagues, although this apparent overlap was not made explicit in Lim et al. We made this determination on the basis of the similarity in the reported provenance of the samples and the genotyping technologies used. Access to these genotype data meant we were able to unambiguously determine the ‘effect’ and ‘other’ allele in the GWAS regression. After processing the summary statistics from this GWAS as described above, we compared the effect direction at each SNP also present in Bronson and colleagues’ data set and found that the direction of effect was consistently reversed in the latter relative to the former (Supplementary Figure 10).

## Data and code availability

The main GWAS data sets, cFDR results, GPS test statistics, and genetic correlation estimates are available at the following Zenodo page: https://zenodo.org/doi/10.5281/zenodo.11929771.

The code used to produce our results is presented in the form of a *snakemake* pipeline [183] at the following GitHub repository: github.com/twillis209/igad_paper_pipeline.

## Notes

### Author Declarations

Norwich Local Research Ethics Committee of Norwich District Health Authority gave ethical approval for this work.

